# How parents judge newborn screening expansion in the genomic era: a theory-informed survey in France from the SeDeN-p3 study

**DOI:** 10.64898/2026.02.22.26346822

**Authors:** Camille Level, Laurence Faivre, Margot Lemaitre, Dominique Salvi, Isabelle Marchetti-Waternaux, Elisabeth Cudry, Emmanuel Simon, Nicolas Bourgon, Alexandra Benachi, Nhut-Thanh Van, Camille Coppola, Christine Binquet, Christel Thauvin-Robinet, Frédéric Huet, Christine Peyron

## Abstract

**Background:** Newborn screening (NBS) has progressively expanded through technological innovations, from tandem mass spectrometry enabling expanded NBS (eNBS) to the prospect of genomic NBS (gNBS). While these developments promise earlier diagnosis and richer information, they also raise concerns regarding actionability, uncertainty, equity and psychosocial impact. As technological feasibility alone does not ensure public confidence, parental perspectives are central to evaluating future expansions. Using acceptability concept as an anticipatory lens, this study assessed parental views on NBS expansion in France, examining its determinants, distinguishing test modalities, and exploring whether genomics raises specific concerns.

**Methods:** A nationwide cross-sectional survey (September 2022–February 2023) included 1,640 parents recruited postpartum in maternity wards and through an online quota panel. Acceptability of eNBS and gNBS, intermediate evaluative components, and sociodemographic characteristics were assessed. Analyses combined descriptive statistics, multivariable regression, and thematic analysis of free-text comments.

**Results:** Support was very high for eNBS (93%) and remained high for gNBS (89%), with genetics mainly shifting responses from complete to partial acceptability. Affective attitude and perceived effectiveness were the strongest predictors of both outcomes, while ethical concerns distinguished assured from conditional support. Most parents prioritised minimising uncertain results, whereas a smaller subgroup accepted greater ambiguity. Foreign-born and single parents reported lower levels of complete acceptability, while health-sector workers and parents with rare-disease experience were more supportive. No independent association with the age of the youngest child was observed.

**Conclusion:** Parental acceptability of eNBS and gNBS is high but nuanced, shaped primarily by anticipated health benefits, emotional orientation and tolerance for uncertainty, with trust and social distance modulating support. As genomic expansion progresses, implementation will require proportionate, culturally adapted information and clear governance, and should be informed by real-world evidence from pilots such as PERIGENOMED.

**Trial registration:** ClinicalTrials.gov, NCT06111456. Last verified: October 2023.

## Introduction

### Newborn screening as an evolving public health programme

Newborn screening (NBS) is widely regarded as one of public health’s major achievements. Since the introduction of Guthrie’s phenylketonuria test and the formulation of the Wilson and Jungner principles in the late 1960s, high-income countries have progressively implemented universal programs to identify treatable disorders early in life. Over recent decades, the scope of NBS has widened with successive technological innovations [1].

Expanded NBS (eNBS), driven by tandem mass spectrometry and related methods, made it possible to screen simultaneously for multiple biochemical disorders and to progressively widen national panels. More recently, the introduction of genome sequencing (GS) in NBS (gNBS) has been presented as a further step change, enabling detection of hundreds of genetic conditions, including those without biochemical markers, and addressing some of the technical limits of standard screening in premature or ill newborns [2–7]. The advantages of such extensions include earlier diagnosis, timely therapies, and the long-term value of population-level data. Yet unresolved challenges (how to define actionability, manage uncertainty, limit false positives, ensure equity, and address psychosocial impacts) have been identified and debated for over a decade across successive waves of NBS expansion, and are now being revisited with added intensity in the genomic context [8–21]. Ongoing real-world pilots provide initial evidence on how these longstanding concerns translate into practice [22]. In this context, understanding how these developments are appraised becomes crucial, as these judgements ultimately determine what is regarded as acceptable within a public health programme. Recent work also underscores that the perspectives of stakeholders are essential to understanding under which conditions such technological shifts can be responsibly implemented [23,24].

### The relevance of test acceptability in NBS

Acceptability offers a relevant framework for analysing how individuals assess future forms of NBS, as it refers to the way they weigh anticipated benefits and burdens across cognitive, affective and normative dimensions [25]. This prospective perspective differs from acceptance, which relates to lived experience after rollout [26], and is particularly pertinent in NBS, where decisions are made upstream of any concrete exposure. Work applying the Theoretical Framework of Acceptability (TFA) has highlighted the central role of perceived appropriateness and shown that judgements depend on how users understand an intervention’s purpose, mechanisms and implications [27]. Within this landscape, parents hold a distinctive position: they are both the recipients of reassurance and the mediators of consent on behalf of their newborn. Studies of standard NBS illustrate that even in systems with near-universal uptake, participation is shaped by values, norms and perceptions of legitimacy, rather than by technical performance alone. Concerns related to pain, alternative medicine, religious beliefs or institutional trust illustrate how these orientations guide parental decision-making [28,29]. These observations echo broader insights from health services research, where acceptability is shown to vary across contexts according to perceived benefits, familiarity with the modality, confidence in handling its informational implications, and understanding of how the intervention works [30–32]. A large body of behavioural research also indicates that evaluative judgements can be sensitive to framing effects, where simple linguistic cues activate heuristics that shift perceptions of benefit and risks [33]. In this perspective, GS may carry symbolic and informational weight, as genetic information is often associated with notions of heredity, identity and long-term implications, and may require higher levels of literacy or interpretive effort. Such representations can shape how parents anticipate potential benefits and burdens.

### International evidence on parental acceptability

In the case of NBS, international studies have explored parental views on eNBS (Canada: [34]; Hong Kong [35]; Netherlands [36,37]; USA [38–40]) or gNBS ((Australia [41–44]; Canada [45,46]; Germany [47]; Slovenia [48]; UK [49]; United Arab Emirates [50]; USA [51–57]), but most have examined these modalities separately and without explicitly comparing how the nature of the test shapes parental evaluations. Synthesising this work, recent reviews describe parental acceptability as insufficiently theorised and often treated descriptively, with many studies relying on hypothetical scenarios or small samples and neglecting the influence of health-system structures and cultural contexts [58,59]. Yet contextual factors consistently appear to shape parental views [18,28]. The French setting, characterised by a strong public health tradition, near-universal participation in standard NBS and structured national governance, provides a particularly informative environment to examine how parents evaluate different modalities of NBS, and whether the informational and symbolic features associated with genomic testing activate specific concerns or reinforce evaluative patterns observed elsewhere [57,60–66].

### The SeDeN project and objectives of this article

This study is part of SeDeN (*Séquençage Dépistage Néonatal*), a multi-component research project examining the conditions under which eNBS and gNBS may be socially acceptable in France. The project comprises four complementary strands: SeDeN-P1, addressing national and international public policies; SeDeN-P2, exploring the views of perinatal and genetics professionals; SeDeN-P3, investigating parental perspectives in the general population and among families affected by rare diseases; and SeDeN-P4, focusing on policymakers and influential stakeholder groups. This article reports findings from SeDeN-P3, specifically the national survey of parents in general population. It pursues two main objectives: first, to identify the determinants of parental acceptability of NBS expansion by explicitly distinguishing the effect of test modality. Second, to characterise patterns of adherence and reservations to determine whether genomic techniques introduce specific concerns or reinforce general evaluative logics observed in eNBS. By comparing both modalities within the same respondents and situating the analysis upstream of a planned national pilot, this study aims to inform future discussions on communication strategies, consent frameworks, and policy options for forthcoming expansions of NBS.

## Materials and methods

### Study design

SeDeN-P3 was a nationwide cross-sectional survey conducted in France between September 2022 and February 2023 using a self-administered questionnaire, administered both online and on paper depending on the recruitment setting. The study followed a convergent mixed-methods design of the “validating quantitative data” type with a concurrent triangulation structure [67], in which quantitative data formed the core and qualitative inputs from open-text responses were used to corroborate and enrich interpretation [68].

This article focuses on parental acceptability of test modalities in NBS. Components addressing criteria for disease inclusion and parental expectations regarding information and consent are reported separately.

### Conceptual framework of acceptability

Acceptability was conceptualised as a multidimensional evaluative judgement combining anticipated benefits, perceived burdens, and alignment with personal values. We relied on the TFA proposed by Sekhon et al., which conceptualises acceptability through multiple components reflecting both cognitive and affective responses to a health intervention [25,69]. This framework was complemented by findings from the international literature on eNBS and gNBS available at the time of questionnaire design and refined through iterative discussions within a multidisciplinary research team (genetics, paediatrics, public health, midwifery, social sciences, and patient organisations).

Fig 1 presents the conceptual framework guiding the study. It organises variables into blocks representing global acceptability outcomes, intermediate components of the acceptability judgement, and respondent characteristics, and outlines the hypothesised links between these elements.

**Fig 1.**
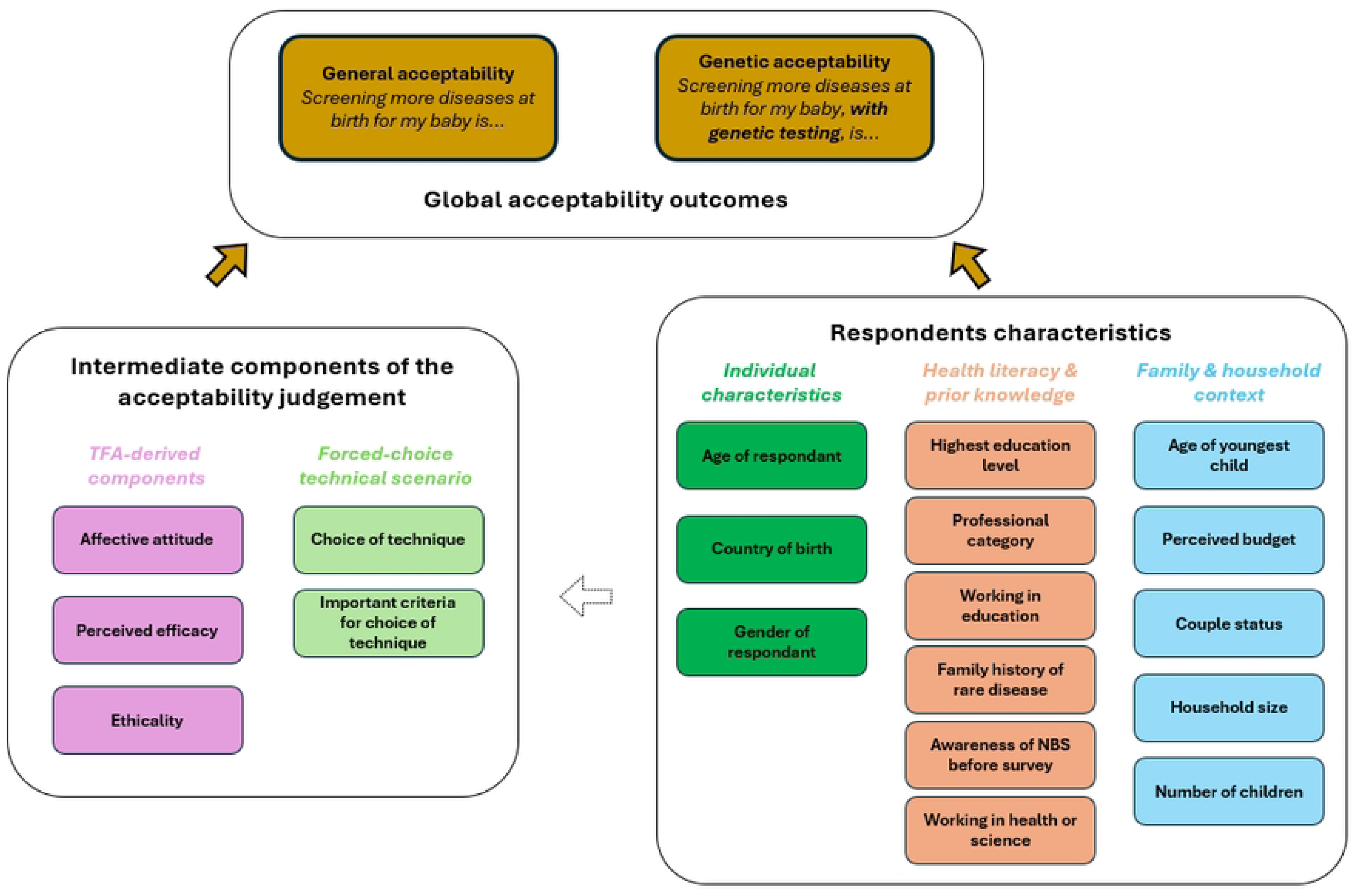
Conceptual framework of acceptability in our study. Fig 1 presents the structure of the key variables in our survey, organised into three interrelated blocks. Arrows indicate the hypothesised influence of these factors on overall and genetic acceptability ratings.

### Survey instrument

Cognitive pre-tests were conducted in maternity wards and with panel participants. Revisions included simplifying medical terminology (e.g. replacing “genome sequencing” with “genetic testing”), shortening item stems to reduce cognitive load, and adjusting response formats for clarity. All core items were mandatory in the electronic version to minimise missing data. The final questionnaire (administered in French and translated into English for publication purposes) is available in S1 File.

#### Global acceptability outcomes

Overall support for eNBS and for gNBS was assessed with two parallel items adapted from Sekhon’s generic acceptability question [69]. Parents rated on a 5-point Likert scale (from *Completely unacceptable* to *Completely acceptable*): (1) “Screening more diseases at birth for my baby is…”, then (2) the same statement specifying that the extension would rely on “a genetic test”. This paired design aimed to isolate the effect of explicitly framing the extension as genetic. An optional free-text box followed the second item.

#### Intermediate components of the acceptability judgement

In addition to the two global acceptability outcomes, the questionnaire captured several intermediate components that clarify the considerations parents may weigh when forming an acceptability judgement, beyond their immediate global rating.

Three components derived from the TFA and relevant to a one-off preventive intervention were included: affective attitude (emotional response to the intervention), perceived effectiveness (extent to which the intervention is expected to achieve its purpose) and ethicality (fit with personal values). Each used a 5-point Likert scale.

A forced-choice technical scenario asked parents to choose between options trading off screening breadth and test precision. Respondents indicated the criterion guiding their choice and could elaborate in free text.

#### Respondent characteristics

Respondent profiles were captured across individual characteristics (age, sex, education, occupation), health literacy and prior knowledge (presence of a rare disease and/or genetic condition in the family, prior personal experience with genetic testing and prior information on NBS), and family and household context (marital status, number of children, financial situation). These variables were treated as potential predictors or stratification factors.

### Participants and recruitment

#### Populations

Two complementary cohorts were recruited to reflect the range of contexts in which parents form judgements about eNBS and gNBS.

Population 1 included parents of newborns ≤ 7 days, recruited exhaustively in four maternity wards between September and November 2022, providing a close approximation of the decision setting at birth. Within each site, recruitment was conducted exhaustively during predefined inclusion periods corresponding to days with on-site recruitment activity, rather than continuously over the entire calendar interval. Sites were selected to maximise diversity in geographic location, facility type and birth volume (S2 Table). Exclusion criteria were neonatal death during recruitment and cognitive or language barriers. A minimum sample size of 385 was calculated using a 95 % confidence level, 5 % margin of error and a conservative 50 % acceptability estimate.

Population 2 included parents of children aged 7 days-36 months, recruited via an online panel (*CPA Études*) between January and February 2023. Quota sampling on region, age and education was implemented using soft quotas, allowing limited deviations to maximise feasibility while approximating the parent population in metropolitan France. The target of 1,155 participants, set to allow comparisons across three child-age groups (0–1, 1–2, 2–3 years), was exceeded to meet quotas (n=1,248).

Combining an exhaustive maternity sample with a broader panel reduced the selection bias typical of voluntary online surveys while capturing potential variation in responses as children age. Analyses were stratified by the age of the youngest child rather than by recruitment modality, to account for potential variation in perceptions as children age.

#### Inclusion criteria

In both populations, parents (biological or otherwise) aged 18 to 50 years for mothers, 60 years for fathers, residing in metropolitan France and not under judicial protection were eligible. These criteria were chosen to ensure legal capacity to consent and to focus on age groups most involved in early childcare and health-related decision-making. Each eligible parent could participate independently, allowing collection of individual-level perceptions within the same household when both partners were available.

### Data analysis strategy

#### Data management

Data were collected via a smartphone-compatible electronic questionnaire. For Population 1, paper forms were also available; these were double entered to minimise transcription errors. The anonymised dataset was stored on the hospital’s encrypted server and was accessible only to authorised study personnel. For optional sociodemographic variables, analyses were conducted using available responses without imputation of missing data.

#### Univariate and bivariate analysis

Univariate analyses were first conducted to describe response distributions for the two global acceptability outcomes and for all intermediate components of the acceptability judgement. Percentages and absolute frequencies are reported for these variables.

Given the absence of full psychometric validation of the TFA-based items and ongoing methodological debate regarding the treatment of 5-point Likert scales [70–73], all items were analysed as ordinal categorical variables. Non-paired associations between global acceptability outcomes, intermediate components and respondent characteristics were tested using Pearson’s χ² or the Fisher-Freeman-Halton exact test when Cochran’s rule was violated. Variables associated with the outcome at p<0.10 were retained for multivariable modelling, consistent with the exploratory aim. Effect sizes were quantified using Cramer’s V (V<0.10 very weak; 0.10–0.19 weak; 0.20–0.29 moderate; ≥0.30 strong).

To assess the impact of explicitly framing screening as genetic, paired differences between eNBS and gNBS ratings were examined using the Wilcoxon signed-rank test (α=0.05). Respondents were classified as showing an unchanged, increased or decreased rating.

#### Multivariable modelling

Predictors of acceptability were examined through separate logistic regression models for eNBS and gNBS. The primary outcome was defined as a single binary dependent variable contrasting positive acceptability (*Somewhat acceptable* or *Completely acceptable*) with non-positive responses (*Somewhat unacceptable* or *Completely unacceptable*). Because most decreases between eNBS and gNBS occurred within the positive range, complementary models distinguishing *Completely acceptable* from *Somewhat acceptable* were also estimated (see Results and S3 Tables). Neutral responses were excluded from modelling due to their heterogeneous interpretation (genuine indecision, ambivalence or uncertainty).

Candidate predictors included TFA-derived components, the criterion guiding the technical trade-off decision, and sociodemographic or experiential characteristics associated with the outcome at p≤0.10. Two modelling strategies were applied: one forcing TFA components into all models, and one unconstrained. Models were screened for multicollinearity using the variance inflation factor (VIF<5) for continuous predictors and Cramer’s V<0.30 for categorical associations, then ranked using Akaike Information Criterion (AIC). The best-fitting specification for each outcome is reported as adjusted odds ratios (OR) with 95 % confidence intervals (CI).

#### Qualitative and mixed-methods integration

Qualitative material from two open-text boxes was analysed thematically by two independent researchers with backgrounds in economics and sociology. Following Creswell’s guidance for convergent mixed methods designs [67], qualitative and quantitative strands were analysed separately and integrated during interpretation. Themes were compared across levels of eNBS and gNBS acceptability and across technical preferences to identify convergences and divergences. Illustrative quotations are presented in the Results when they clarify or contextualise quantitative patterns.

Open-text comments were mapped onto a two-dimensional matrix defined by respondents paired eNBS and gNBS acceptability ratings. Each comment was represented as a bubble positioned according to the combination of ratings, with bubble size reflecting the number of respondents sharing the same rating configuration. This visual mapping was used as an exploratory tool to support the identification of qualitative profiles.

#### Software

Analyses were conducted using Microsoft Excel (version 2506) and R via Rstudio (version 4.4.2.) for quantitative analysis and NVivo for qualitative data management and thematic analysis. Data cleaning support was provided by ADN Soft via the HARMONIE platform.

## Results

### Recruitment and respondent characteristics

A total of 1,775 parents initiated the questionnaire, of whom 1,640 completed it, yielding an overall completion rate of 92.4% (Fig 2), Among completed questionnaires, 392 were collected in maternity wards (Population 1) and 1,248 via the online panel (Population 2). In Population 1, the estimated coverage rate was 58.6% (414/707), reflecting the proportion of eligible parents approached during predefined inclusion periods who initiated the survey (S2 Table). In Population 2, 1,361 of 9,217 individuals who accessed the survey initiated it, yielding a 14.8% conversion rate. Respondent characteristics are presented in Table 1.

**Fig 2.**
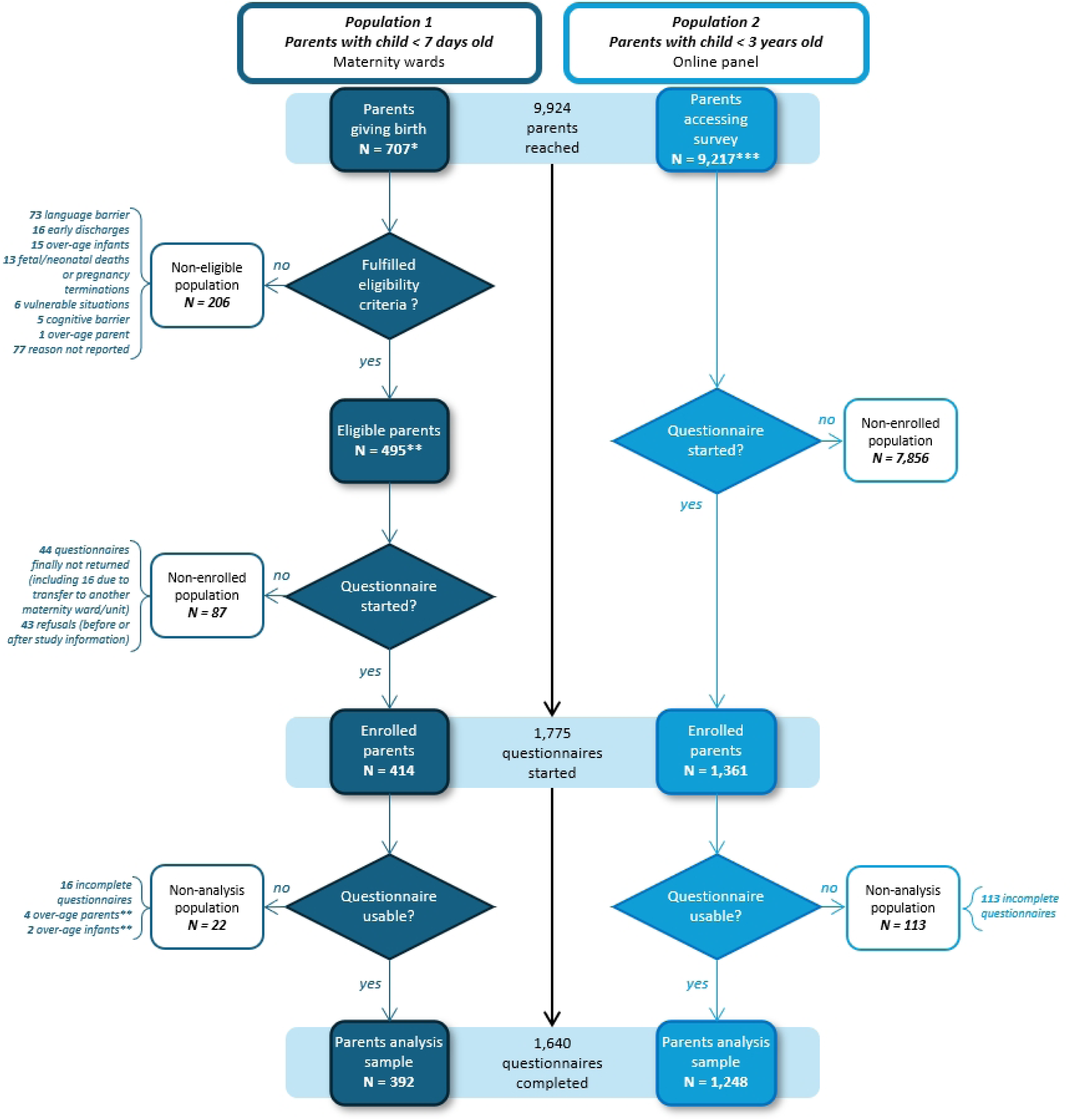
Participant flow for the two recruitment populations. * Based on national estimates from the 2021 DREES survey on childcare arrangements (https://drees.solidarites-sante.gouv.fr/sources-outils-et-enquetes/lenquete-modes-de-garde-et-daccueil-des-jeunes-enfants), 89.7% of parents of children under age 3 in metropolitan France live in a two-parent household. Applying this proportion to the 373 deliveries recorded during the recruitment period in Population 1, we estimate that approximately 707 parents could theoretically have been reached to complete the questionnaire. This estimate accounts for both two-parent households (two potential respondents per delivery) and single-parent households (one respondent) and was used to define the upper bound of potential respondents for coverage calculations. ** Of the 22 non-analysed cases, 6 were found post hoc to be ineligible (4 outside age range, 2 with a child older than 7 days) and were therefore excluded from the denominator of eligible participants to ensure consistency with inclusion criteria. *** In Population 2, 9,217 individuals accessed the survey page. The estimated parent population is not directly known. Among them, 1,361 answered the first substantive question, representing the starting point for inclusion. The gap with initial clicks (n=7,854) reflects early exits, quota ineligibility, or disengagement before consent or first question.

**Table 1.**
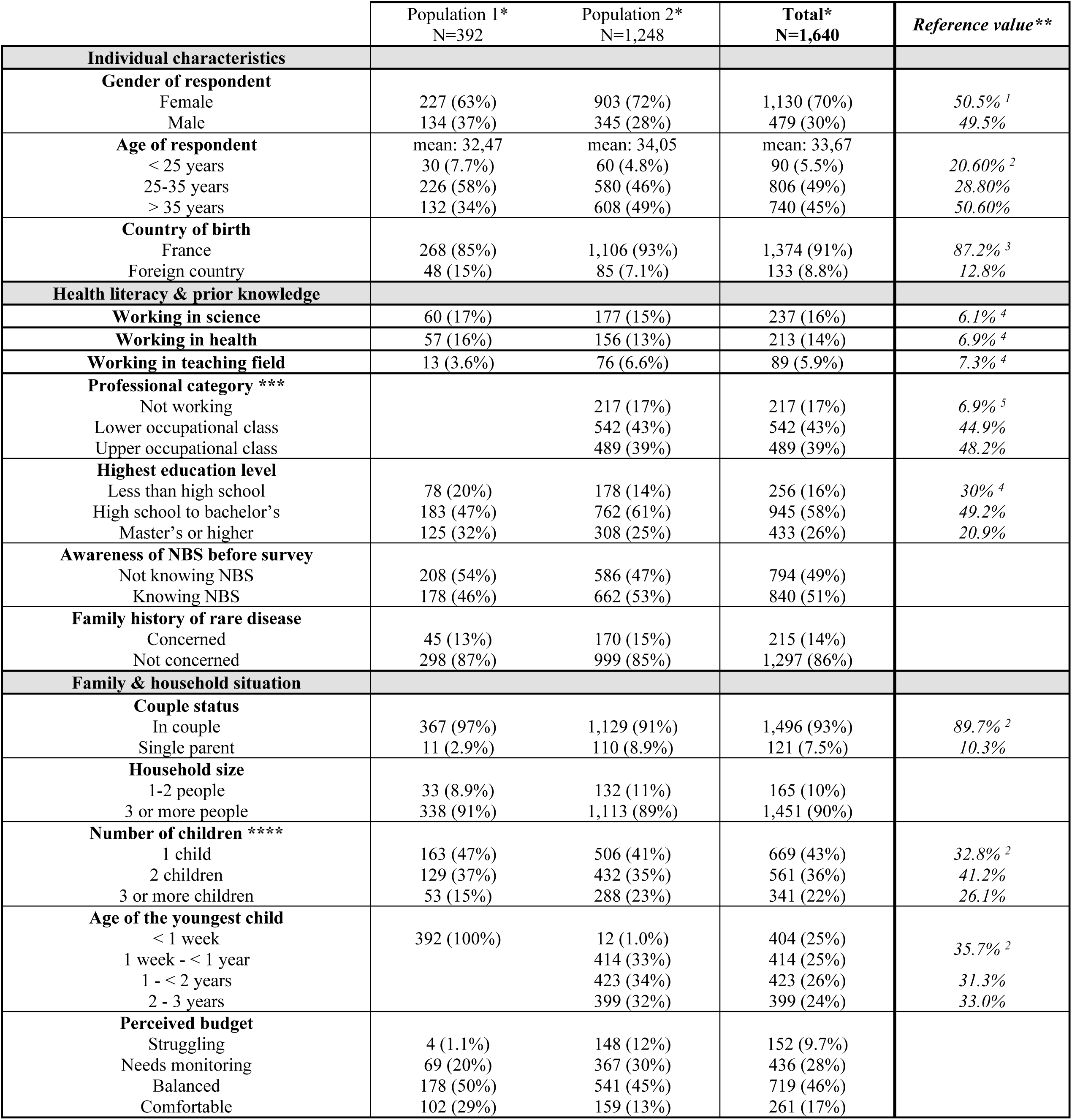
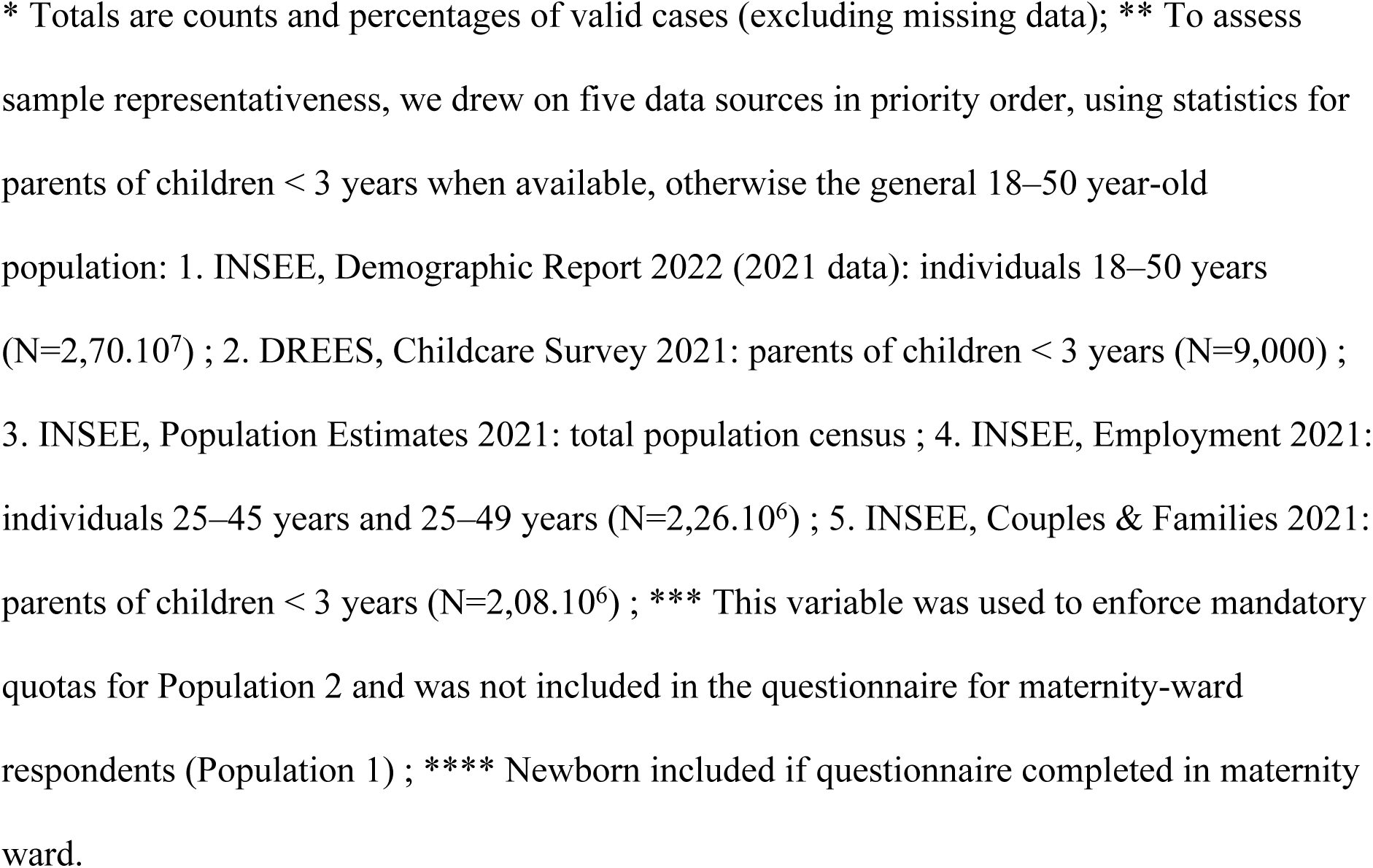
Respondents’ characteristics, by recruitment site and population group, with reference values.

### Global acceptability outcomes and opinion shifts

93.2% (n=1,528) of respondents rated the idea of *Screening more diseases at birth for their child* as *Somewhat acceptable* or *Completely acceptable*. When the genetic dimension was explicitly introduced (by specifying *using a genetic test, DNA analysis*), this general level slightly declined to 89.3% (n=1,463).

Among the 1,631 participants who provided responses to both paired acceptability items, 64.2% (n=1,047) gave identical ratings, including 48.6% (n=793) who consistently chose *Completely acceptable* (Fig 3). Most changes observed in the remaining third of respondents were modest, with 27.1% (n=443) moving downwards by one category (most often from *Completely acceptable* to *Somewhat acceptable*, n=327; 20.0%) and only 4.6% (n=76) moved from a positive to a negative category. Conversely, 8.6% (n=141) shifted upwards. The distribution of paired responses differed significantly (Wilcoxon signed-rank test, V=126,261.5, p<0.0001), indicating that explicit mention of genetic testing modified acceptability ratings for a subset of parents, while overall support remained high.

**Fig. 3.**
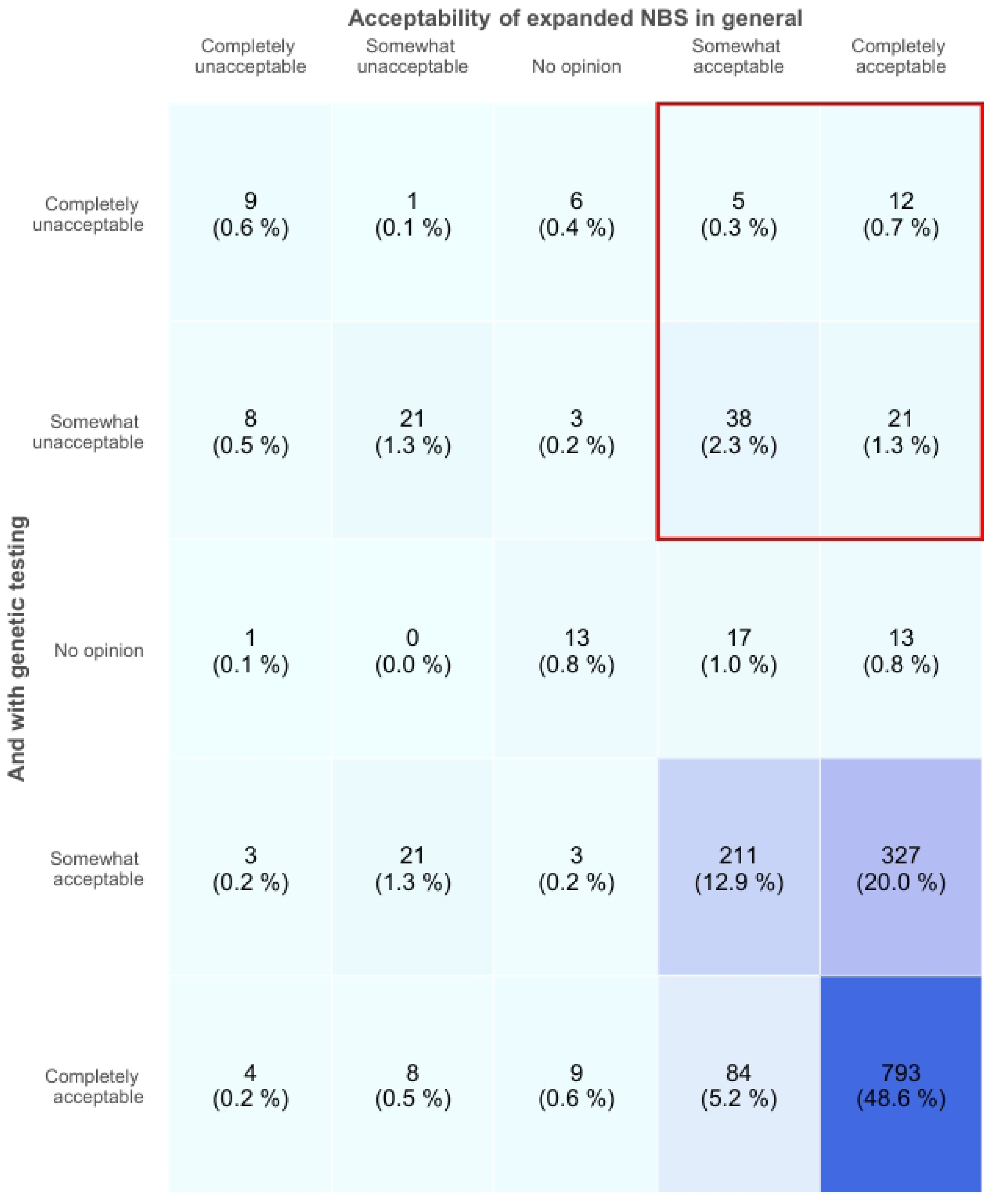
Cross-distribution of responses to general and genomic expanded newborn screening acceptability. Heatmap of paired responses (N=1,631) showing high overall acceptability for both scenarios. Most participants maintained a positive opinion. The red box highlights downward transitions from a previously supportive stance (n=76; 4.6%).

Acceptability outcomes were examined across age groups of the youngest child (< 1 week; 1 week–< 1 year; 1–< 2 years; 2–3 years). Overall support for eNBS remained high across all age groups. Positive acceptability (*Somewhat acceptable* or *Completely acceptable*) exceeded 90% in each group for eNBS and remained above 85% when genetic testing was explicitly mentioned. Differences across age groups were modest and followed similar patterns of slight downward shifts when genetics was introduced (Tables 2a and 2b).

**Table 2.**
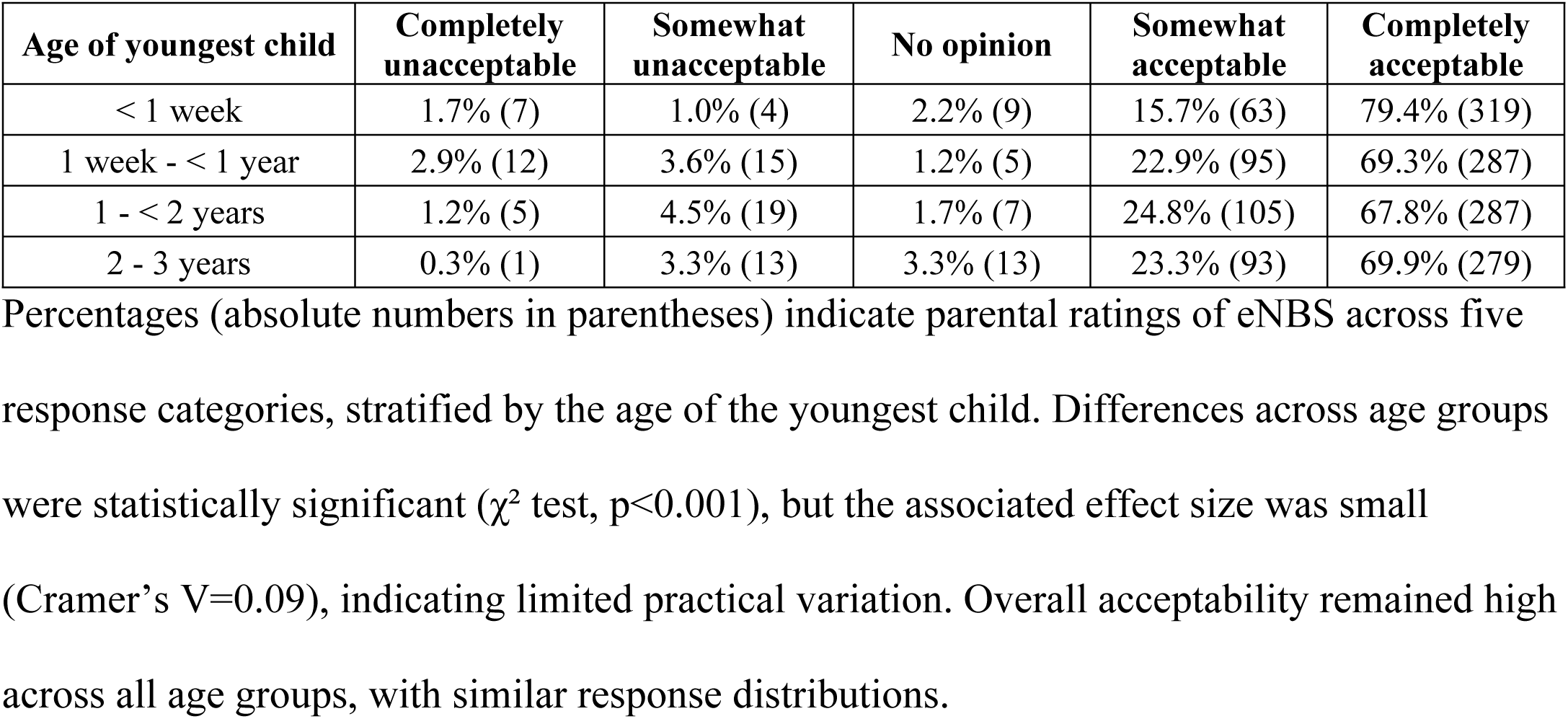
Global acceptability of eNBS and gNBS by age of the youngest child. (2a) Global acceptability of eNBS by age of the youngest child

| Age of youngest child | Completely unacceptable | Somewhat unacceptable | No opinion | Somewhat acceptable | Completely acceptable |
| --- | --- | --- | --- | --- | --- |
| < 1 week | 1.7% (7) | 1.0% (4) | 2.2% (9) | 15.7% (63) | 79.4% (319) |
| 1 week - < 1 year | 2.9% (12) | 3.6% (15) | 1.2% (5) | 22.9% (95) | 69.3% (287) |
| 1 - < 2 years | 1.2% (5) | 4.5% (19) | 1.7% (7) | 24.8% (105) | 67.8% (287) |
| 2 - 3 years | 0.3% (1) | 3.3% (13) | 3.3% (13) | 23.3% (93) | 69.9% (279) |

Percentages (absolute numbers in parentheses) indicate parental ratings of eNBS across five response categories, stratified by the age of the youngest child. Differences across age groups were statistically significant ( $\chi^2$ test, $p<0.001$ ), but the associated effect size was small (Cramer's V=0.09), indicating limited practical variation. Overall acceptability remained high across all age groups, with similar response distributions.

(2b) Global acceptability of gNBS by age of the youngest child.

| Age of youngest child | Completely unacceptable | Somewhat unacceptable | No opinion | Somewhat acceptable | Completely acceptable |
| --- | --- | --- | --- | --- | --- |
| < 1 week | 3.0% (12) | 3.5% (14) | 5.0% (20) | 34.0% (135) | 54.4% (216) |
| 1 week - < 1 year | 1.7% (7) | 6.5% (27) | 2.7% (11) | 37.2% (154) | 51.9% (215) |
| 1 - < 2 years | 2.4% (10) | 7.1% (30) | 0.9% (4) | 32.6% (138) | 57.0% (241) |
| 2 – 3 years | 1.0% (4) | 5.3% (21) | 2.5% (10) | 34.6% (138) | 56.6% (226) |

Percentages (absolute numbers in parentheses) indicate parental ratings of gNBS across five response categories, stratified by the age of the youngest child. Differences across age groups were statistically significant ( $\chi^2$ test, $p=0.014$ ), with a small effect size (Cramer's $V=0.07$ ). Despite this statistical difference, acceptability patterns were comparable across age groups, and support for gNBS remained high regardless of the age of the youngest child.

### Intermediate components of the acceptability judgement

For affective attitude, 88.4% (n=1,441/1,640) agreed or strongly agreed with the statement *I would like more diseases to be screened at birth for my baby*. For perceived effectiveness, 92.4% of respondents (n=1,507) agreed or strongly agreed that *Screening more diseases at birth for [their] baby can improve [its] health*. Regarding ethicality, 62.2% (n=1,014) disagreed or strongly disagreed that *Screening more diseases at birth for their baby raises moral or ethical concerns*, whereas 31.4% (n=515) agreed or strongly agreed, indicating that nearly one third of parents perceived ethical questioning associated with eNBS.

When presented with three hypothetical screening strategies - Technique A (many diseases, high uncertainty risk), Technique B (moderate number of diseases and risk), and Technique C (few diseases, low risk) - 43.9% (n=713) of parents selected Technique C, while 36.5% (n=594) selected Technique B and 19.6% (n=318) Technique A. Across all respondents, 55.0% (n=892) reported that minimising uncertainty was the main criterion guiding their choice. Cross-tabulation confirmed this pattern (Table 3): among those preferring Technique C, 79.7% (n=568) explicitly prioritised reducing uncertainty, whereas parents selecting Technique A more frequently cited the number of diseases (52.5%; n=167). Technique B showed a balanced distribution between the two criteria.

**Table 3.**
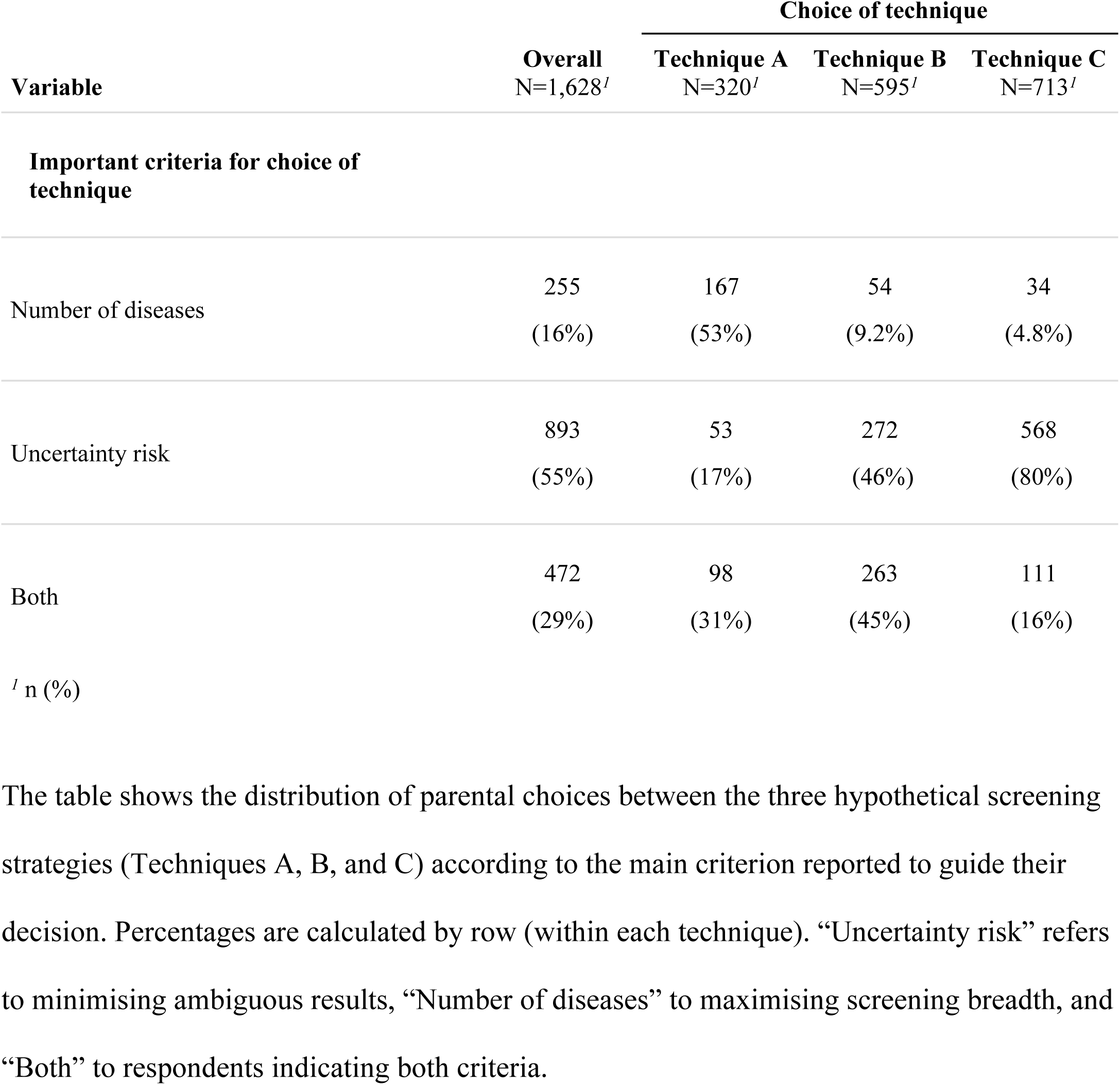
Distribution of parental screening strategy choices by decision criterion.

| Variable | Overall<br>N=1,628<sup>/</sup> | Choice of technique |  |  |
| --- | --- | --- | --- | --- |
|  |  | Technique A<br>N=320<sup>/</sup> | Technique B<br>N=595<sup>/</sup> | Technique C<br>N=713<sup>/</sup> |
| Important criteria for choice of technique |  |  |  |  |
| Number of diseases | 255 | 167 | 54 | 34 |
|  | (16%) | (53%) | (9.2%) | (4.8%) |
| Uncertainty risk | 893 | 53 | 272 | 568 |
|  | (55%) | (17%) | (46%) | (80%) |
| Both | 472 | 98 | 263 | 111 |
|  | (29%) | (31%) | (45%) | (16%) |

<sup>1</sup> n (%)
The table shows the distribution of parental choices between the three hypothetical screening strategies (Techniques A, B, and C) according to the main criterion reported to guide their decision. Percentages are calculated by row (within each technique). “Uncertainty risk” refers to minimising ambiguous results, “Number of diseases” to maximising screening breadth, and “Both” to respondents indicating both criteria.

Among respondents who indicated that minimizing uncertainty was the main factor guiding their choice of technique, open-ended responses strongly emphasized the risks of parental anxiety, stress, and potential psychological harm associated with uncertain or unreliable results. Typical comments include: “*The results must be very reliable, otherwise they are too anxiety-inducing*” and “*False results can have a very serious impact on the child’s health but also on the parents’ mental health.*”. Respondents prioritizing the number of diseases screened tended to justify their choices by emphasizing the importance of maximizing early detection and clinical benefit and often acknowledging the need for confirmatory testing in case of positive results. Finally, those who selected both criteria typically advocated for a balance between broad screening and test reliability, frequently recommending combined or sequential testing approaches to ensure both comprehensive and reliable results. Full coded verbatims are provided in S4 Table.

### Multivariable regression models

#### Predictors of positive acceptability (Somewhat/Completely acceptable vs Somewhat/Completely unacceptable)

In the general acceptability model (Fig 4a), a positive perceived efficacy was the strongest predictor (OR 17.7, 95% CI 7.6–42.0, p<0.001), as was higher affective attitude (OR 9.5, 95% CI 4.2-21.3, p<0.001). Respondents born outside France and single parents were less likely to support expansion (resp. OR 0.3, 95% CI 0.1–0.9, p=0.020; OR 0.3, 95% CI 0.1–0.9, p=0.018) whereas employment in the health sector was associated with greater support (OR 6.3, 95% CI 1.5–44.9, p=0.028).

**Fig. 4.**
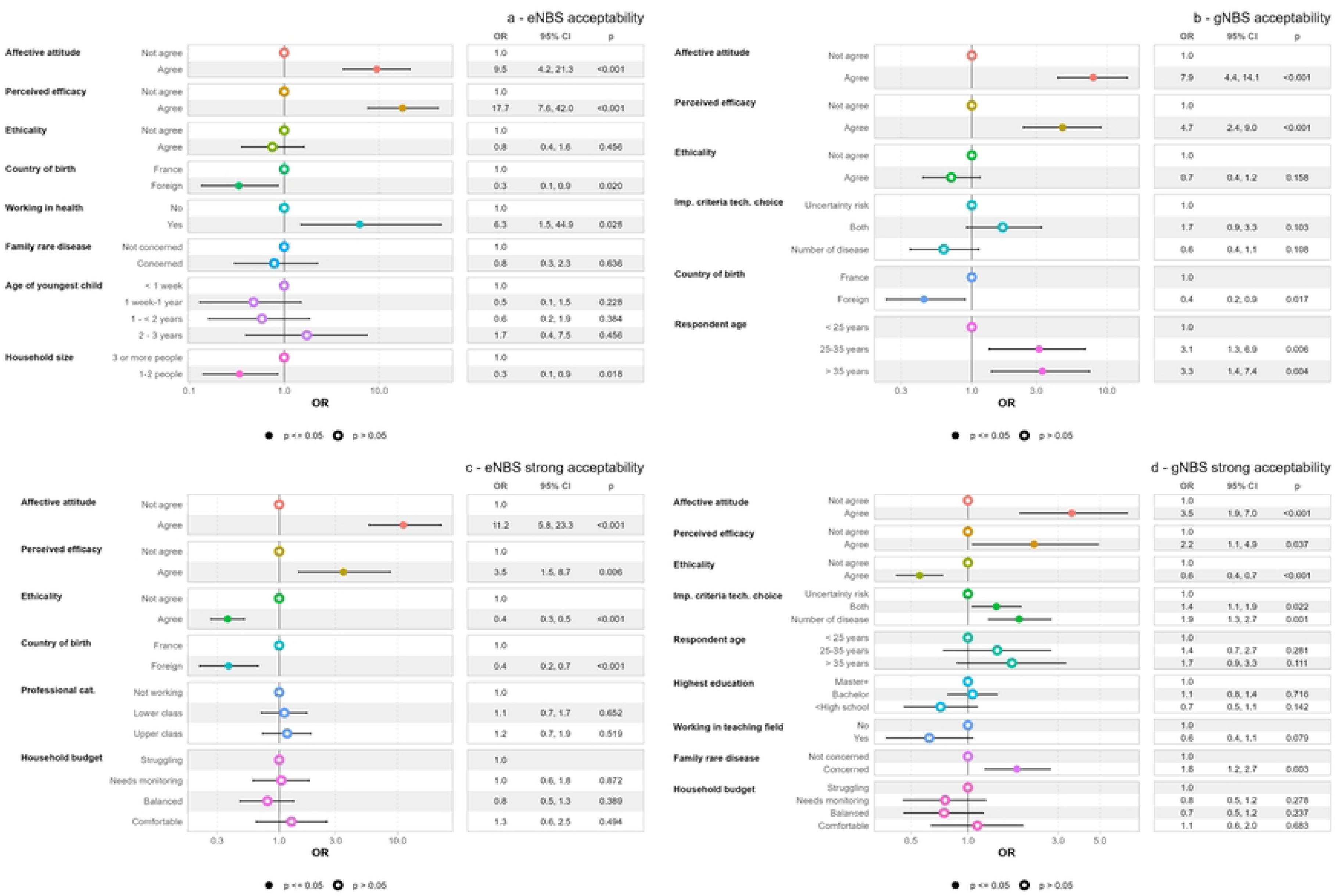
Multivariable regression forest plots for eNBS and gNBS acceptability. For each model, odds ratios (OR) and their 95% confidence intervals (CI) are shown for each explanatory variable (adjusted for all covariates included in the model). Filled circles indicate statistically significant associations (p≤0.05); open circles represent non-significant results. The table to the right of each plot displays the exact OR, 95% CI, and p-value for each modality. Reference categories (OR=1) are shown at the top of each variable block. Models shown: 4a: General acceptability (*Completely acceptable/Somewhat acceptable* vs. *Completely unacceptable*/*Somewhat unacceptable*); 4b: Genomic acceptability (*Completely acceptable/Somewhat acceptable* vs. *Completely unacceptable*/*Somewhat unacceptable*); 4c: General strong acceptability (*Completely acceptable* vs. *Somewhat acceptable*); 4d: Genomic strong acceptability (*Completely acceptable* vs. *Somewhat acceptable*).

In the model assessing acceptability when genetics was explicitly mentioned (Fig 4b), affective attitude (OR 7.9, 95% CI 4.4–14.1, p<0.001) and perceived efficacy (OR 4.7, 95% CI 2.4–9.0, p<0.001) remained significant predictors. Foreign-born respondents again showed lower support (OR 0.4, 95% CI 0.2–0.9, p=0.017). Compared with respondents aged 18–24 years (reference category), those aged 25–35 years (OR 3.1, 95% CI 1.3–6.9, p=0.006) and those older than 35 years (OR 3.3, 95% CI 1.4–7.4, p=0.004) were more likely to support genomic expansion. The criterion guiding choices in the technical trade-off scenario (minimising uncertainty vs maximising screening breadth) was not associated with overall positive acceptability for either eNBS or gNBS, after adjustment.

#### Predictors of strong acceptability (Completely acceptable vs Somewhat acceptable)

In complementary models (Fig 4c–d), several associations persisted. Strong acceptability for eNBS (Fig 4c) was more frequent among respondents with higher affective attitude (OR 11.2, 95% CI 5.8–23.3, p<0.001) and perceived effectiveness (OR 3.5, 95% CI 1.5–8.7, p=0.006) scores, and less frequent among those expressing ethical concerns (OR 0.4, 95% CI 0.3–0.5, p<0.001). Foreign-born status was linked to more moderate support (OR 0.4, 95% CI 0.2–0.7, p<0.001).

For strong acceptability of gNBS (Fig 4d), affective attitude (OR 3.5, 95 % CI 1.9–7.0, p<0.001) and perceived efficacy (OR 2.2, 95% CI 1.1–4.9, p=0.037) remained positive predictors, while ethical concerns again lowered odds (OR 0.6, 95 % CI 0.4–0.7, p<0.001). Respondents with a family history of rare disease were more likely to be fully supportive (OR 1.8, 95 % CI 1.2–2.7, p=0.003). Compared with the majority who reported being guided by minimizing uncertainty in their choice of screening strategy, respondents who placed greater weight on maximizing the number of diseases and less on uncertainty, were more likely to consider gNBS as *Completely acceptable* (Both: OR 1.4, 95 % CI 1.1–1.9, p=0.022; Number of diseases: OR 1.9, 95 % CI 1.3–2.7, p=0.001).

Across all multivariable models, the age of the youngest child was not independently associated with acceptability outcomes after adjustment, indicating that the modest age-related differences observed in descriptive analyses were not sustained once other factors were considered.

### Qualitative material and mapping of parental profiles

A total of 81 respondents (4.9%) provided open-text comments (S5a Table). These were positioned on a bubble matrix according to each respondent’s combined eNBS and gNBS ratings (Fig 5). Cross-tabulation is presented in S5b Table. Five qualitative profiles emerged from this mapping. The largest group (Zone 1, n=48) consisted of parents who rated both scenarios as highly acceptable. Their comments were benefit-centred, highlighting earlier treatment, informed decision-making, valuing knowledge and anticipatory planning, often framed as an “obvious necessity,” with occasional references to the importance of non-invasive procedures. A smaller set of respondents (Zone 2, n=3) expressed an even stronger endorsement when genetics was specified; for them, the prospect of genomic analysis heightened rather than altered an already favourable view. Another sizeable group (Zone 3, n=18) also maintained positive ratings across both scenarios, but their support was explicitly conditional. Their narratives emphasized requirements such as non-invasiveness, restricted use of samples, strong data confidentiality and clear legal safeguards, often invoking broader principles of proportionality and ethical oversight. By contrast, respondents in Zone 4 (n=8) were favourable to eNBS in general but shifted downward when the genetic dimension was introduced. Their comments pointed to concerns about privacy, fears of eugenics and a perception that existing safeguards were insufficient. Finally, a smaller set of respondents (Zone 5, n=4) consistently rejected both scenarios, invoking ethical or existential arguments such as the sanctity of life, the rejection of medical over-reach and the fear of eugenics. Mentions of anticipated psychological burden (n=4, highlighted in orange in Fig 5) were distributed across profiles and did not cluster within a specific acceptability pattern.

**Fig. 5.**
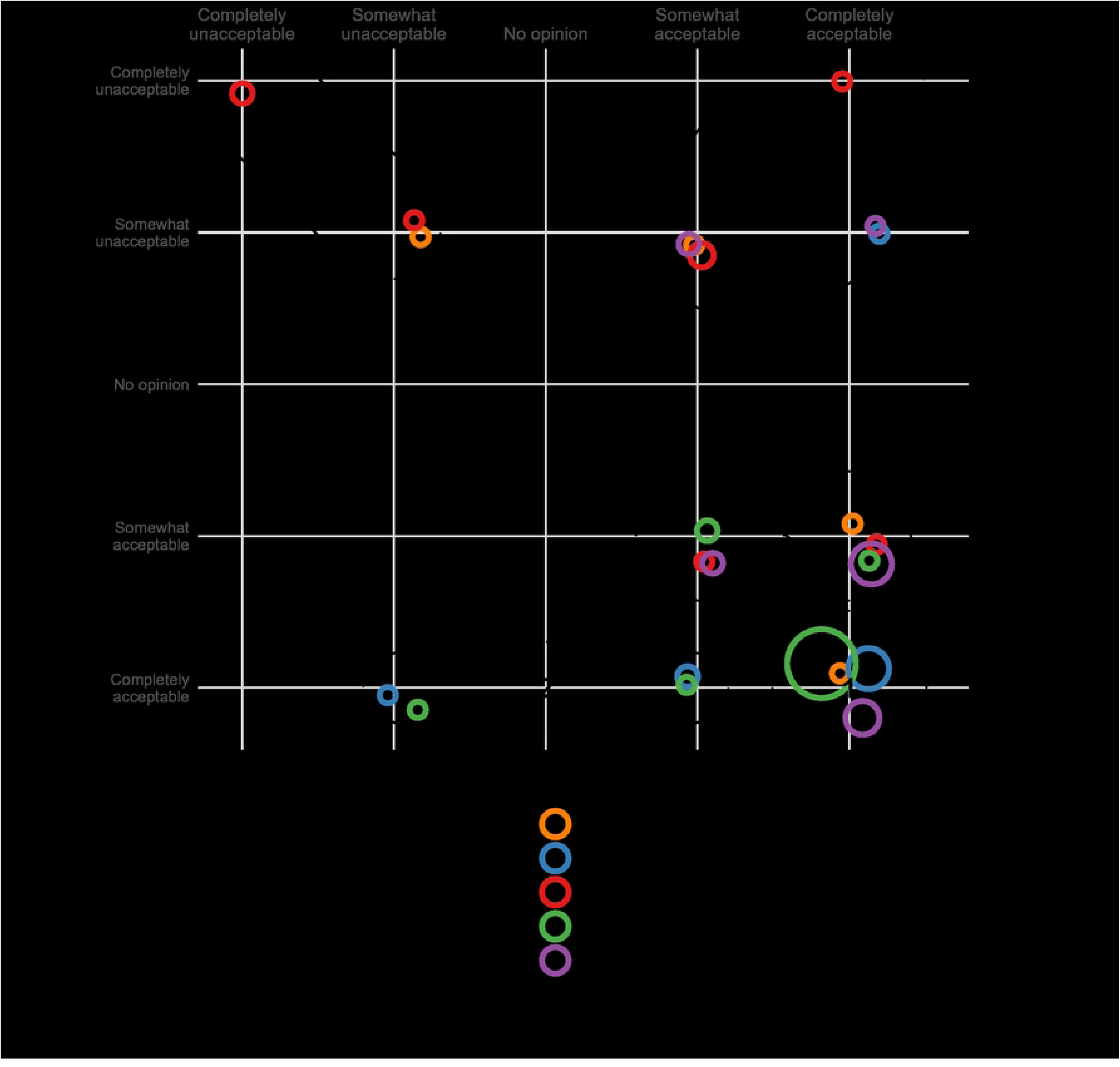
Mapping of parental comments by shift in acceptability between general and genomic NBS. Bubble plot mapping parental verbatim comments (n=81) onto paired acceptability ratings of expanded (x-axis) and genomic newborn screening (y-axis). Each bubble represents one theme comment, positioned according to the respondent’s global acceptability scores. Bubble size is proportional to the number of respondents sharing the same position, while bubble colour indicates the main thematic category of the associated comment: (1) information and consent (orange), (2) perceived health benefits (blue), (3) concerns about uncertainty or psychosocial impact (red), (4) ethical or value-based considerations (green), and (5) broader views on science, society, and governance (purple). Dotted ellipses delineate the five shift zones (1-5). The grid of the plot defines distinct zones of movement: along the diagonal (stable acceptability between expanded and genomic versions), below it (decrease in acceptability when genetics is introduced), and above it (increase in acceptability).

## Discussion

### Main findings and contribution

The results presented here provide empirical evidence on parental acceptability of eNBS and gNBS in France, and show how test modality, particularly the explicit mention of genomic testing, shapes parental judgement.

Overall support for eNBS was extremely high (93%), and acceptability remained close to this level (89%) when the extension was explicitly framed as genetic. Most shifts occurred within positive evaluations, from *Completely acceptable* to *Somewhat acceptable*, and transitions to refusal were rare. Across modalities, three components adapted from the TFA (affective attitude, perceived effectiveness and ethicality) were the strongest correlates of parental judgement. Positive emotional orientation toward screening and confidence in its effectiveness were robust predictors of both eNBS and gNBS acceptability, whereas ethical concerns mainly distinguished unconditional from more conditional forms of support. Technical preferences added further nuance: most parents prioritised reliability and the minimisation of uncertain results, while a smaller subgroup accepted greater ambiguity and expressed stronger support for gNBS. Sociodemographic and experiential characteristics (foreign-born status, single parenthood, health-sector employment, family history of rare disease) shaped these patterns but remained secondary to perceptual and value-based components. Because parental judgements about NBS may vary with time since birth, we examined acceptability outcomes according to the age of the youngest child rather than by recruitment setting. Descriptive analyses showed modest age-related differences, with consistently high levels of support for both eNBS and gNBS across all age groups and similar patterns of slightly lower acceptability when genetic testing was explicitly mentioned. Although statistically significant, these differences were associated with small effect sizes. Importantly, age of the youngest child was not independently associated with acceptability outcomes in multivariable models. This suggests that parental evaluations of eNBS and gNBS are shaped less by the immediacy of the postnatal context than by broader evaluative dimensions such as perceived benefits, affective responses and ethical considerations, supporting interpretation across populations.

This study extends the literature by providing large-scale, theory-informed evidence on parental views, addressing gaps highlighted in recent reviews [24,58,59]. With 1,640 parents, including a substantial number of postpartum participants, it complements earlier work conducted mainly in North America, Australia and Canada, which relied on smaller or convenience samples.

### Contextualisation within prior research

#### Global parental support for NBS expansion

Our findings align with international prospective surveys showing strong support for standard NBS expansions and high, but slightly lower, support for genomic extensions (S6 Table). In the French context, this decline in acceptability when genetic testing is explicitly mentioned appears more modest than that reported in several international studies. In the United States, 78.7% of parents in a control arm versus 71.0% in a GS arm strongly agreed that every newborn should undergo screening [74]. In Australia, interest decreased from 99% for standard NBS to 77% for gNBS [75]. In Canada, willingness to participate was 94.4% for current NBS versus 79.6% for genome-based screening [76]. Comparable gradients appear in eNBS surveys, where parents commonly endorsed broad panels, including less treatable or untreatable childhood-onset disorders [34,36–40]. For gNBS, international evidence shows high interest, though explicit reference to “genetics”, late-onset conditions, or uncertain results tends to moderate enthusiasm [41,43,44,51,53,56]. These patterns are consistent with framing effects described in behavioural economics [33]: explicitly labelling an extension as “genetic” appears to trigger simple heuristics that temper, without reversing, support. In that sense, parental judgements reflect trade-offs in which perceived benefits are weighed against potential burdens rather than a binary “yes/no” stance, and unpacking the underlying components helps explain how very high overall acceptability can coexist with more moderate endorsements of genomic modalities.

#### Why parents want screening: anticipated gains, reassurance and actionability

In our study, two TFA dimensions clearly emerged as the strongest drivers of parental acceptability: affective attitude (the wish for one’s child to benefit from screening) and perceived effectiveness (the belief that eNBS or gNBS would improve a child’s health). Regression analyses indicated that affective attitude accounted for a substantial share of variance in overall acceptability, echoing the central role of behavioural intention in the Theory of Reasoned Action, where attitudes toward a behaviour strongly shape intention independently of detailed cost–benefit calculations [77,78] and its application to eNBS by Paquin et al. [39]. Across studies, anticipated health benefits consistently emerge as the main justification for supporting eNBS [34,41,47,50,53,54,74,79]. Our qualitative material reinforced this pattern: parents most often referred to earlier diagnosis, improved prognosis and the reassurance associated with “knowing”, echoes found across prior studies [34,56,57,80]. Several parents also emphasised the anticipatory value of information - for planning, emotional preparedness or family organisation - an orientation similarly highlighted in the literature. Together, these observations show that information is valued not only for its clinical utility but also for its capacity to reduce uncertainty and support decision-making. Taken together, these findings suggest that parental judgements rest first on a positive emotional orientation and a belief in effectiveness, consistent across both eNBS and gNBS. From this standpoint, genomic techniques in NBS seem evaluated primarily as extensions of an already valued programme rather than as a radical break.

#### Uncertainty, reliability and the primacy of “doing no harm”

Uncertainty aversion was a defining feature of parental judgement. When asked to choose between breadth and precision, most parents preferred strategies that minimised uncertain or inconclusive results, even at the cost of screening fewer conditions. This pattern is consistent with earlier eNBS and gNBS studies that highlight hesitations around late-onset conditions, lower test accuracy or limited treatment options [34,38,42,47]. Open-text comments in our study directly linked uncertain results to parental anxiety, psychological burden and potential disruption of family life.

A smaller subgroup showed higher tolerance for uncertain or ambiguous results and was more likely to judge gNBS *Completely acceptable*, indicating heterogeneity in how parents weigh uncertainty against potential benefits. International studies report similar divides over late-onset conditions, lower test accuracy, penetrance thresholds and the risk of parental anxiety [42,47,81]. These gradients are consistent with behavioural concepts such as loss aversion and with the notion of absorptive capacity, i.e. the ability to take in, process and act on complex information [82].

Although relatively uncommon in our data, some parents raised concerns about anxiety, loss of carefree parenting or potential changes in the parent–child relationship in the event of uncertain results. Such fears have been more extensively documented in gNBS contexts [47,54] and, earlier, in eNBS research on ambiguous metabolic findings and false positives [9,34,83,84]. These observations suggest that, for a subset of families, informational gains may be outweighed by anticipated psychosocial costs.

Non-invasiveness also formed part of this calculus. Several parents explicitly mentioned discomfort linked to heel-prick procedures, noting that pain or mishandling could make screening feel disproportionate to its purpose. Similar reactions have been reported elsewhere [29,82]. At the same time, many parents in our study acknowledged that the health benefits of eNBS outweighed procedural inconvenience, a balance also observed by Berrios et al. (2020) and Wang et al. (2022).

Within the TFA, these themes map onto burden and intervention coherence and underline that acceptability depends not only on perceived benefit but also on the effort and discomfort associated with participation. Taken together, they indicate that even when affective attitude and perceived effectiveness remain strong, procedural and informational burdens must be actively managed if acceptability is to be sustained. In this context, genomic testing appears acceptable not because it is novel, but because parents judge that its informational yield is, or is not, proportionate to its psychological and organisational implications.

#### Ethical concerns, trust and social distance

Ethical concerns rarely translated into outright refusal but tended to distinguish reserved from unconditional support. Fears of eugenics, data misuse and over-medicalisation, which appeared in our qualitative material, are consistent with gNBS literature emphasising consent, privacy, discrimination risks and data governance [41,50,53,57,74,80]. Similar themes have also been reported in eNBS, particularly around blood sample handling, pain and data use [28,35].

Patterns of trust and social distance were also apparent. Lower acceptability among parents born outside France and among single parents echoes findings on the role of social reinforcement, trust and normative influence. Paquin et al. (2016) showed that perceived support from partners or relatives was a strong predictor of intention to participate in eNBS, helping to explain why parents with less social backing may appear more cautious. Van der Pal and colleagues identified religious beliefs and reliance on alternative medicine as frequent reasons for refusing current NBS, while Wang et al. documented strong endorsement of gNBS among Latina mothers in the United States when information was adapted to their linguistic and cultural context.

Conversely, greater support among health-sector workers or parents with rare-disease experience mirrors studies linking familiarity, personal relevance and improved literacy to more favourable attitudes [36,48,56,85–88]. For many of these parents, the wish to avoid diagnostic odysseys and to secure earlier intervention appears to tilt the balance in favour of expansion.

Taken together, these findings suggest that acceptability is not solely a psychological response to a test but also a relational judgement anchored in trust and situated within broader social and cultural frames. While affective orientation and perceived effectiveness remain the main levers of support, sociodemographic and experiential factors modulate how parents weigh benefits against risks. In this sense, gNBS operates as a test of trust: what is at stake is not only the technology itself, but also confidence in the institutions that organise, govern and explain it. This has direct implications for implementation and governance, underscoring the importance of transparent procedures, inclusive deliberation and tailored communication strategies to support equitable participation.

### Implications for implementation and governance

#### Information strategies that enable, rather than burden, decision-making

Parents valued information that was understandable and proportionate. Communication should therefore avoid both oversimplification and overload. Presenting the concrete health benefits for the child and the anticipated value of early detection upfront aligns with the strongest observed drivers of acceptability and may reduce the initial hesitancy associated with the label “genetic”, a pattern consistent with behavioural framing effects. At the same time, parents expected uncertainties to be acknowledged rather than minimised. Adapting materials to different levels of health literacy, and providing linguistically and culturally appropriate versions, appears necessary to reduce informational inequalities, particularly for families who feel more distant from the health system.

#### Governance, transparency and the integration of public values

Findings suggest that sustaining parental acceptability will depend at least as much on governance as on technical performance. Ethical and institutional dimensions such as data protection, clarity on analytic scope, procedures for residual bloodspots and accessible channels for queries or appeals seem to function as structural determinants of trust rather than peripheral safeguards. For this reason, implementation would benefit from making these elements visible, consistent and proportionate. In parallel, governance frameworks may need to integrate parental values more explicitly. Involving parent and patient representatives in the development of information materials, consent procedures and data-governance policies could help ensure that expansion proceeds with restraint, fairness and clarity of purpose. Embedding routine evaluation of parental experience within programme oversight may further support accountability and alignment with families lived realities. Evidence from prior work shows that actively involving parents in the design and governance of eNBS or gNBS can strengthen both ethical legitimacy and program sustainability [57,60–66].

#### Ensuring operational proportionality as genomic expansion progresses

Parents tended to support genomic expansion when they perceived that benefits outweighed psychological and organisational burdens. Maintaining this balance requires demonstrating that expanded pathways remain manageable for families and feasible for professionals. Priorities include minimising additional steps, ensuring non-invasive procedures, providing rapid confirmatory testing, and offering structured support for the communication and interpretation of uncertain results. In this perspective, operational coherence, rather than technological novelty, is what sustains confidence and acceptability.

### Limitations and perspectives

This study has several methodological limitations that affect how the findings should be interpreted. First, the design was prospective: parents evaluated hypothetical scenarios rather than making real-world decisions. Behavioural research shows that hypothetical framing may overestimate uptake because it removes logistical, emotional and temporal pressures experienced in practice [89]. At the same time, Paquin et al. showed that stated intentions can align closely with enrolment decisions when options are offered, supporting the relevance of prospective acceptability as an anticipatory, though not predictive, tool. Evidence from gNBS pilots illustrates this gap. Participation has ranged from very low in BabySeq, where disclosure included adult-onset and uncertain findings [80,90], to very high in Belgium, where only treatable childhood conditions were offered [91]. Intermediate uptake in GUARDIAN and Baby Beyond Hearing similarly reflects strong influence of contextual factors such as timing, parental fatigue, additional procedures or absence of explanation [81,92]. Despite these differences, the core drivers identified prospectively here (anticipated benefit, informational value and sensitivity to uncertainty) consistently appear in real-world settings, supporting the relevance of anticipatory acceptability as a complementary but not predictive tool. Second, selection bias must be acknowledged. The dual recruitment strategy (postpartum participants and an online quota panel) broadened coverage but also introduced limitations.

Two data-collection modalities were used (paper in maternity wards and online questionnaires), and while descriptive analyses showed no major divergence in global acceptability levels, these samples are not strictly comparable. Recruitment required French literacy and digital access, which likely under-represented non-French-speaking parents and those facing acute social vulnerability. Online-panel recruitment may also introduce self-selection biases related to digital literacy or interest in health topics. Although widely used and validated in health research, such panels can still be influenced by unobservable characteristics linked to participation [93]. This likely resulted in underestimation of sociocultural gradients, consistent with international pilots showing lower participation among non-native speakers or families less anchored in the health system [28,56]. These factors may contribute to an underestimation of cultural or community-specific barriers and reinforce the need for targeted inclusion strategies in future work. Third, our measurement strategy only captured part of the multidimensional structure of acceptability. Within the TFA, affective attitude and perceived effectiveness were robustly measured, but constructs such as burden, opportunity costs or intervention coherence were not included, the latter being less salient in the French universal-care setting. While coherent with current practice, this limits external validity, as cost has emerged as a key driver in other settings [40,44,48,50]. Some concerns (non-invasiveness, confidentiality, legal safeguards) were only captured in free-text comments, preventing their integration into regression models. Although the TFA has been predominantly used in implementation research in low- and middle-income settings [94–97], our results point to the relevance of its core components in a universal-care context. We do not interpret this as a validation of the framework, but as an indication that emotional, cognitive and value-based judgements may play a stable role across different health-system configurations. Further psychometric work would nevertheless be necessary to confirm the transferability of these constructs to NBS settings. Fourth, the study did not examine how acceptability evolves over time, particularly across the transition from pregnancy to the postpartum period, where absorptive capacity and psychological fatigue may alter decision-making. Real-world pilots show that contextual barriers (additional blood draws, lack of explanation or partner disagreement) shape decisions in ways that hypothetical assessments cannot fully capture [81,98]. Longitudinal designs are therefore needed to understand how initial intentions are reframed once gNBS is offered. Finally, several essential dimensions of screening governance were beyond the scope of this article. Criteria for inclusion, storage and reanalysis rules, and secondary uses of data, all central to public and professional debates, were not examined here. Preferences regarding treatability, onset, actionability and acceptable scope will be addressed in a separate article. Divergences between parental and professional rationalities, already documented elsewhere [85,86], warrant dedicated comparative analysis to clarify how different actors define the boundaries of “acceptable” expansion.

Taken together, these limitations highlight three directions for future work. First, expanding the sociocultural scope of research. Including non-French-speaking parents, integrating culturally adapted materials, and measuring constructs such as trust, acculturation and health-system familiarity would reduce current blind spots and enable a more accurate assessment of sociocultural gradients in acceptability. Second, clarifying governance arrangements and inclusion criteria. Parents value anticipatory information even when treatments are lacking, while simultaneously acknowledging the psychological risks of ambiguous findings. Understanding how parents and professionals weigh these competing rationalities can help structure frameworks for proportionate and socially legitimate expansion. Third, strengthening the bridge between prospective and real-world evidence. Combining anticipatory surveys with longitudinal follow-up in implementation pilots, including PERIGENOMED in France, would make it possible to analyse how parents navigate uncertainty, procedural burdens and psychosocial impacts once screening becomes part of lived experience. More broadly, positioning acceptability as both an ex-ante evaluative lens and a bridge to implementation science may help ensure that eNBS and gNBS are developed in ways that remain clinically robust, socially legitimate and sustainable.

## Conclusions

This study shows that parental acceptability of eNBS and gNBS is high yet shaped by identifiable evaluative logics. Judgements rested primarily on anticipated benefits for the child and on a positive emotional orientation toward screening, while tolerance for uncertainty and concerns about governance differentiated assured from conditional support. Although explicit reference to genetics moderated enthusiasm, it did not reverse overall endorsement, suggesting that gNBS is largely interpreted as an extension of an already valued programme. Sociodemographic and experiential factors modulated these orientations, indicating that trust and cultural distance remain important for implementation. Taken together, these findings underline that the success of any expansion will depend not only on technical performance but also on proportionate, culturally adapted communication and transparent governance.

Prospective acceptability therefore offers a useful anticipatory lens, but real-world evidence from forthcoming pilots such as PERIGENOMED will be essential for guiding responsible and socially legitimate adoption.

## Data Availability

The data underlying the results of this study contain sensitive personal information collected from parents in a postpartum context and cannot be shared publicly due to ethical and legal restrictions, in accordance with the approval granted by the Ethics Committee for Research of the University of Burgundy–Franche-Comté. An anonymised dataset is available from the corresponding author for researchers who meet the criteria for access to confidential data, subject to approval by the relevant ethics committee and the signing of a data use agreement.

## List of abbreviations

NBS: Newborn screening
eNBS: Expanded newborn screening
gNBS: Genomic newborn screening
TFA: Theoretical Framework of Acceptability
GS: Genome sequencing
SeDeN: Séquençage et Dépistage Néonatal (national survey and research project)
CI: Confidence interval
OR: Odds ratio
AIC: Akaike Information Criterion
VIF: Variance inflation factor

## Declarations

## Ethics approval and consent to participate

The protocol, data collection tools, and information sheets for families were approved by the Ethics Committee for Research of the University of Burgundy-Franche-Comté on September 8, 2022 (S7 File). Participation was voluntary. All participants received written information describing the study objectives, procedures, data collected, and their rights. In accordance with French regulations for non-interventional research, participation was based on informed non-opposition, documented by completion of the questionnaire after receipt of the information notice. For Population 1, information was provided on-site by trained research staff during the maternity stay, using a written information and non-opposition notice. For Population 2, the same information was provided on the first page of the online questionnaire. Data were collected anonymously and analysed in accordance with applicable data protection regulations.

## Availability of data and materials

The data underlying the results of this study contain sensitive personal information collected from parents in a postpartum context and cannot be shared publicly due to ethical and legal restrictions, in accordance with the approval granted by the Ethics Committee for Research of the University of Burgundy–Franche-Comté.

An anonymised dataset is available from the corresponding author for researchers who meet the criteria for access to confidential data, subject to approval by the relevant ethics committee and the signing of a data use agreement.

## Authors’ contributions

CL (Camille Level), LF (Laurence Faivre), ML (Margot Lemaitre), DS (Dominique Salvi), IM (Isabelle Marchetti-Waternaux), EC (Elisabeth Cudry), ES (Emmanuel Simon), NB (Nicolas Bourgon), AB (Alexandra Benachi), TV (Nhut-Thanh Van), CC (Camille Coppola), CB (Christine Binquet), CTR (Christel Thauvin-Robinet), FH (Frédéric Huet), CP (Christine Peyron)

CL conceived and designed the study, coordinated recruitment, supervised data management, conducted statistical and qualitative analyses, and drafted the first manuscript. CP and LF contributed to study design, defined objectives and hypotheses, supported interpretation of results, and provided substantial revisions. IM, EC, CB, CTR and FH contributed to the development of the study protocol and questionnaire. DS supported recruitment organisation, data collection logistics, and data preparation. ES, NB, AB, TV and CC facilitated recruitment in maternity wards, presented the study to parents and administered questionnaires. ML contributed to qualitative analysis. All authors critically reviewed the manuscript and approved the final version.

## Acknowledgements

The authors gratefully acknowledge the contribution of all parents who participated in the survey. We also thank the clinical teams in the participating maternity wards for their support with recruitment and questionnaire administration. We are grateful to ADN Soft for their assistance with data management and analysis.

## Supporting information

**Supplementary material S1. Survey Instrument.** Full questionnaire administered to parents, including information sheets, consent procedures, sociodemographic items, acceptability measures, and evaluative components. Yellow highlights indicate online programming and skip logic; page breaks are marked.

**Supplementary material S2. Site-level recruitment characteristics and completion rates (Population 1).** Characteristics of participating maternity wards, recruitment periods, inclusion and non-inclusion reasons, and questionnaire completion metrics for the maternity-based recruitment (Population 1). * All multiple births recorded during the recruitment periods were twin births.

**Supplementary material S3. Individual shifts in acceptability with mention of genetic testing and additional regression analyses.** Cross-tabulations showing changes in individual acceptability judgments when genetic testing is explicitly mentioned, by sociodemographic characteristics, health literacy, and family situation. All observed associations were of small magnitude, with Cramer’s V values ranging from 0.06 to 0.12. Supplementary Table provides detailed cross-tabulations, cell counts, percentages, and exact p-values for all outcomes. This table summarizes the associations between respondent sociodemographic characteristics (rows) and the three main outcome variables (columns): general acceptability of expanded newborn screening, acceptability of expanded newborn screening with genetics, and individual opinion shift with mention of genetic testing. For each characteristic, cell values are presented as: n (% within characteristic) - that is, count and row percentage for each response category. Percentages are calculated excluding missing data. Totals for each subgroup are displayed in the first column. Total counts vary across variables due to missing data, as sociodemographic characteristics were not mandatory in the questionnaire. p-values (and Cramer’s V, if p < 0.1) are reported for each comparison, based on either Pearson’s Chi-squared or Fisher-Freeman-Halton (FFH) test according to expected cell counts (Cochran’s rule, simulation = 2000). Abbreviations: NBS: newborn screening, n: count, %: row percentage, Cramer’s V: effect size measure for categorical associations, ‡: FFH, *p < 0.1, **p < 0.05, ***p < 0.01, ***p < 0.001. This supplementary file also presents the best-fitting logistic regression models for parental acceptability outcomes, based on explanatory variables recoded as binary (agree or important vs not agree or not important). Models are reported for acceptability of eNBS and gNBS, using alternative outcome codings. Two specifications are shown: models including core acceptability components derived from the Theoretical Framework of Acceptability (affective attitude, perceived effectiveness, ethicality), and models estimated without this constraint. Model comparison relied on Akaike Information Criterion (AIC), variance inflation factors, and sample size. These analyses assess model robustness and the stability of associations across specifications.

**Supplementary material S4. Themes expressed by parents regarding in their choice of screening technique.** This supplementary table presents free-text comments organised by thematic categories (rows) and by the screening technique criteria judged important by respondents (columns). This layout allows comparison of thematic patterns across technical preference profiles.

**Supplementary material S5. Thematic analysis of comments on acceptability. S5a: Catalogue of verbatims classified by macro-theme and sub-theme, with definitions (n=81 unique verbatims)** Colours match the palette used in Figure 4. Sub-themes are listed in decreasing order of frequency. When a comment addressed several sub-themes, it was repeated and counted in each relevant category. **S5b: Cross-tabulation of general and genomic acceptability among respondents who provided a free-text comment (n = 81 unique verbatims)** Rows represent acceptability with genetic testing; columns represent acceptability in general. Each cell shows the count followed by the percentage of the comment subgroup.

**Supplementary material S6. Selection of published studies on parental attitudes toward newborn screening.** Overview of selected published studies examining parental attitudes toward newborn screening and its expansion, including study design, population, and main findings.

**Supplementary material S7. Ethics approval for the SeDeN-p3 study.** Favourable opinion issued by the Ethics Committee for Research of the University of Burgundy–Franche-Comté (CER UBFC) for the SeDeN-p3 study, dated 8 September 2022 (reference number CERUBFC-2022-04-27-015), including the original approval document and its English translation.

## References

1. Loeber JG, Platis D, Zetterström RH, Almashanu S, Boemer F, Bonham JR, et al. Neonatal Screening in Europe Revisited: An ISNS Perspective on the Current State and Developments Since 2010. Int J Neonatal Screen. 2021;7: 15. doi:10.3390/ijns7010015

2. Remec ZI, Trebusak Podkrajsek K, Repic Lampret B, Kovac J, Groselj U, Tesovnik T, et al. Next-Generation Sequencing in Newborn Screening: A Review of Current State. Front Genet. 2021;12. doi:10.3389/fgene.2021.662254

3. Stark Z, Scott RH. Genomic newborn screening for rare diseases. Nat Rev Genet. 2023;24: 755–766. doi:10.1038/s41576-023-00621-w

4. Vears DF, Savulescu J, Christodoulou J, Wall M, Newson AJ. Are We Ready for Whole Population Genomic Sequencing of Asymptomatic Newborns? Pharmacogenomics Pers Med. 2023;16: 681–691. doi:10.2147/PGPM.S376083

5. Willig LK, Petrikin JE, Smith LD, Saunders CJ, Thiffault I, Miller NA, et al. Whole-genome sequencing for identification of Mendelian disorders in critically ill infants: a retrospective analysis of diagnostic and clinical findings. Lancet Respir Med. 2015;3: 377–387. doi:10.1016/S2213-2600(15)00139-3

6. Bodian DL, Klein E, Iyer RK, Wong WSW, Kothiyal P, Stauffer D, et al. Utility of whole-genome sequencing for detection of newborn screening disorders in a population cohort of 1,696 neonates. Genet Med. 2016;18: 221–230. doi:10.1038/gim.2015.111

7. Wang X, Wang Y-Y, Hong D-Y, Zhang Z-L, Li Y-H, Yang P-Y, et al. Combined genetic screening and traditional biochemical screening to optimize newborn screening systems. Clin Chim Acta. 2022;528: 44–51. doi:10.1016/j.cca.2022.01.015

8. Grob R, Roberts S, Timmermans S. Families’ Experiences with Newborn Screening: A Critical Source of Evidence. Hastings Cent Rep. 2018;48: S29–S31. doi:10.1002/hast.881

9. Buchbinder M, Timmermans S. Newborn screening for metabolic disorders: parental perceptions of the initial communication of results. Clin Pediatr (Phila). 2012;51: 739–744. doi:10.1177/0009922812446011

10. Esquerda M, Palau F, Lorenzo D, Cambra FJ, Bofarull M, Cusi V, et al. Ethical Questions Concerning Newborn Genetic Screening. Clin Genet. n/a. doi:10.1111/cge.13828

11. Friedman JM, Cornel MC, Goldenberg AJ, Lister KJ, Sénécal K, Vears DF, et al. Genomic newborn screening: public health policy considerations and recommendations. BMC Med Genomics. 2017;10: 9. doi:10.1186/s12920-017-0247-4

12. Spiekerkoetter U, Bick D, Scott R, Hopkins H, Krones T, Gross ES, et al. Genomic newborn screening: Are we entering a new era of screening? J Inherit Metab Dis. 2023;46: 778–795. doi:10.1002/jimd.12650

13. Baple EL, Scott RH, Banka S, Buchanan J, Fish L, Wynn S, et al. Exploring the benefits, harms and costs of genomic newborn screening for rare diseases. Nat Med. 2024;30: 1823–1825. doi:10.1038/s41591-024-03055-x

14. Berg JS, Agrawal PB, Bailey DB, Beggs AH, Brenner SE, Brower AM, et al. Newborn Sequencing in Genomic Medicine and Public Health. Pediatrics. 2017;139: e20162252. doi:10.1542/peds.2016-2252

15. Reinstein E. Challenges of using next generation sequencing in newborn screening. Genet Res. 2015;97: e21. doi:10.1017/S0016672315000178

16. Friedman JM. Implementing genomic newborn screening as an effective public health intervention: sidestepping the hype and criticism. Npj Genomic Med. 2024;9: 70. doi:10.1038/s41525-024-00451-7

17. Tarini BA, Goldenberg AJ. Ethical issues with newborn screening in the genomics era. Annu Rev Genomics Hum Genet. 2012;13: 381–393. doi:10.1146/annurev-genom-090711-163741

18. Bombard Y, Miller FA, Hayeems RZ, Avard D, Knoppers BM. Reconsidering reproductive benefit through newborn screening: a systematic review of guidelines on preconception, prenatal and newborn screening. Eur J Hum Genet EJHG. 2010;18: 751–760. doi:10.1038/ejhg.2010.13

19. Meade C, Bonhomme NF. Newborn screening: adapting to advancements in whole-genome sequencing. Genet Test Mol Biomark. 2014;18: 597–598. doi:10.1089/gtmb.2014.1558

20. Knoppers BM, Sénécal K, Borry P, Avard D. Whole-Genome Sequencing in Newborn Screening Programs. Sci Transl Med. 2014 [cited 23 Dec 2021]. doi:10.1126/scitranslmed.3008494

21. Howard HC, Knoppers BM, Cornel MC, Wright Clayton E, Sénécal K, Borry P, et al. Whole-genome sequencing in newborn screening? A statement on the continued importance of targeted approaches in newborn screening programmes. Eur J Hum Genet EJHG. 2015;23: 1593–1600. doi:10.1038/ejhg.2014.289

22. Magnifico G, Artuso I, Benvenuti S. A systematic review of real-world applications of genome sequencing for newborn screening. Rare Dis Orphan Drugs J. 2023;2: N/A-N/A. doi:10.20517/rdodj.2023.17

23. Goldenberg AJ, Lloyd-Puryear M, Brosco JP, Therrell B, Bush L, Berry S, et al. Including ELSI research questions in newborn screening pilot studies. Genet Med. 2019;21: 525–533. doi:10.1038/s41436-018-0101-x

24. Downie L, Halliday J, Lewis S, Amor DJ. Principles of Genomic Newborn Screening Programs: A Systematic Review. JAMA Netw Open. 2021;4: e2114336. doi:10.1001/jamanetworkopen.2021.14336

25. Sekhon M, Cartwright M, Francis JJ. Acceptability of healthcare interventions: an overview of reviews and development of a theoretical framework. BMC Health Serv Res. 2017;17: 88. doi:10.1186/s12913-017-2031-8

26. Bobillier-Chaumon M-É, Dubois M. L’adoption des technologies en situation professionnelle : quelles articulations possibles entre acceptabilité et acceptation ? Trav Hum. 2009;72: 355–382. doi:10.3917/th.724.0355

27. Bartlett YK, Kenning C, Crosland J, Newhouse N, Miles LM, Williams V, et al. Understanding acceptability in the context of text messages to encourage medication adherence in people with type 2 diabetes. BMC Health Serv Res. 2021;21: 608. doi:10.1186/s12913-021-06663-2

28. Pal SM van der, Wins S, Klapwijk JE, Dijk T van, Kater-Kuipers A, Ploeg CPB van der, et al. Parents’ views on accepting, declining, and expanding newborn bloodspot screening. PLOS ONE. 2022;17: e0272585. doi:10.1371/journal.pone.0272585

29. Etchegary H, Nicholls SG, Tessier L, Simmonds C, Potter BK, Brehaut JC, et al. Consent for newborn screening: parents’ and health-care professionals’ experiences of consent in practice. Eur J Hum Genet EJHG. 2016;24: 1530–1534. doi:10.1038/ejhg.2016.55

30. Pongiglione B, Carrone F, Angelucci A, Mazziotti G, Compagni A. Patient characteristics associated with the acceptability of teleconsultation: a retrospective study of osteoporotic patients post-COVID-19. BMC Health Serv Res. 2023;23: 230. doi:10.1186/s12913-023-09224-x

31. Vatrasresth J, Prapaisilp P, Sukrong M, Sinthuchai N, Karroon P, Maitreechit D, et al. Acceptability of telemedicine for follow up after contraceptive implant initiation at an obstetrics and gynecologic training center. BMC Health Serv Res. 2023;23: 817. doi:10.1186/s12913-023-09816-7

32. Sciamanna CN, Diaz J, Myne P. Patient attitudes toward using computers to improve health services delivery. BMC Health Serv Res. 2002;2: 19. doi:10.1186/1472-6963-2-19

33. Kahneman D, Tversky A. Prospect Theory: An Analysis of Decision under Risk. Econometrica. 1979;47: 263–291. doi:10.2307/1914185

34. Hayeems RZ, Miller FA, Bombard Y, Avard D, Carroll J, Wilson B, et al. Expectations and values about expanded newborn screening: a public engagement study. Health Expect Int J Public Particip Health Care Health Policy. 2015;18: 419–429. doi:10.1111/hex.12047

35. Mak CM, Lam CW, Law CY, Siu WK, Kwong LLT, Chan KL, et al. Parental attitudes on expanded newborn screening in Hong Kong. Public Health. 2012;126: 954–959. doi:10.1016/j.puhe.2012.08.002

36. Plass AMC, El CG van, Pieters T, Cornel MC. Neonatal Screening for Treatable and Untreatable Disorders: Prospective Parents’ Opinions. Pediatrics. 2010;125: e99–e106. doi:10.1542/peds.2009-0269

37. van Dijk T, Kater A, Jansen M, Dondorp WJ, Blom M, Kemp S, et al. Expanding Neonatal Bloodspot Screening: A Multi-Stakeholder Perspective. Front Pediatr. 2021;9: 1078. doi:10.3389/fped.2021.706394

38. Lipstein EA, Nabi E, Perrin JM, Luff D, Browning MF, Kuhlthau KA. Parents’ decision-making in newborn screening: opinions, choices, and information needs. Pediatrics. 2010;126: 696–704. doi:10.1542/peds.2010-0217

39. Paquin RS, Peay HL, Gehtland LM, Lewis MA, Bailey DB. Parental intentions to enroll children in a voluntary expanded newborn screening program. Soc Sci Med. 2016;166: 17–24. doi:10.1016/j.socscimed.2016.07.036

40. DeLuca JM. Public Attitudes Toward Expanded Newborn Screening. J Pediatr Nurs. 2018;38: e19–e23. doi:10.1016/j.pedn.2017.10.002

41. Lynch F, Best S, Gaff C, Downie L, Archibald AD, Gyngell C, et al. Australian Public Perspectives on Genomic Newborn Screening: Risks, Benefits, and Preferences for Implementation. Int J Neonatal Screen. 2024;10: 6. doi:10.3390/ijns10010006

42. Lynch F, Best S, Gaff C, Downie L, Archibald AD, Gyngell C, et al. Australian public perspectives on genomic newborn screening: which conditions should be included? Hum Genomics. 2024;18: 45. doi:10.1186/s40246-024-00611-x

43. Tutty E, Archibald AD, Downie L, Gaff C, Lunke S, Vears DF, et al. Key informant perspectives on implementing genomic newborn screening: a qualitative study guided by the Action, Actor, Context, Target, Time framework. Eur J Hum Genet. 2024;32: 1599–1605. doi:10.1038/s41431-024-01650-7

44. Peters R, Best S, Lynch F, Vears DF, Downie L, Archibald AD, et al. Public preferences for the value and implementation of genomic newborn screening: Insights from two discrete choice experiments in Australia. Am J Hum Genet. 2025;112: 1515–1527. doi:10.1016/j.ajhg.2025.05.001

45. Etchegary H, Dicks E, Green J, Hodgkinson K, Pullman D, Parfrey P. Interest in Newborn Genetic Testing: A Survey of Prospective Parents and the General Public. Genet Test Mol Biomark. 2012;16: 353–358. doi:10.1089/gtmb.2011.0221

46. Etchegary H, Dicks E, Hodgkinson K, Pullman D, Green J, Parfey P. Public Attitudes About Genetic Testing in the Newborn Period. J Obstet Gynecol Neonatal Nurs. 2012;41: 191–200. doi:10.1111/j.1552-6909.2012.01341.x

47. Doll ES, Mahal J, Alex K, Lerch SP, Kölker S, Schaaf CP, et al. Tension between the need for certainty and numerous uncertainties-A focus group study on various perspectives on a potential genomic newborn screening program in Germany. J Genet Couns. 2025;34: e70004. doi:10.1002/jgc4.70004

48. Prosenc B, Cizek Sajko M, Kavsek G, Herzog M, Peterlin B. Perception of genomic newborn screening among peripartum mothers. Eur J Hum Genet. 2024;32: 163–170. doi:10.1038/s41431-023-01497-4

49. van Mil H. WGS for newborn screening. 2021.

50. Elhadi YAM, Alkatheeri M, Alktifan M, Alhammadi F, Sultan T, Alqumboz YMA, et al. Parents’ perspectives on expanded newborn genomic screening in Abu Dhabi, United Arab Emirates. Hum Genomics. 2025;19: 63. doi:10.1186/s40246-025-00766-1

51. Waisbren SE, Bäck DK, Liu C, Kalia SS, Ringer SA, Holm IA, et al. In the Early Postpartum Period, Parents are Interested in Newborn Genomic Testing. Genet Med Off J Am Coll Med Genet. 2015;17: 501–504. doi:10.1038/gim.2014.139

52. Dodson DS, Goldenberg AJ, Davis MM, Singer DC, Tarini BA. Parent and Public Interest in Whole Genome Sequencing. Public Health Genomics. 2015;18: 151–159. doi:10.1159/000375115

53. Goldenberg AJ, Dodson DS, Davis MM, Tarini BA. Parents’ interest in whole-genome sequencing of newborns. Genet Med. 2014;16: 78–84. doi:10.1038/gim.2013.76

54. Moultrie RR, Paquin R, Rini C, Roche MI, Berg JS, Powell CM, et al. Parental Views on Newborn Next Generation Sequencing: Implications for Decision Support. Matern Child Health J. 2020;24: 856–864. doi:10.1007/s10995-020-02953-z

55. Gold NB, Omorodion JO, del Rosario MC, Rivera-Cruz G, Hsu CY, Ziniel SI, et al. Preferences of parents from diverse backgrounds on genomic screening of apparently healthy newborns. J Genet Couns. n/a. doi:10.1002/jgc4.1994

56. Wang H, Page R, Lopez D, Arkatkar S, Young C, Martinez D, et al. Pregnant Latinas’ views of adopting exome sequencing into newborn screening: A qualitative study. Genet Med. 2022;24: 1644–1652. doi:10.1016/j.gim.2022.04.012

57. Joseph G, Chen F, Harris-Wai J, Puck JM, Young C, Koenig BA. Parental Views on Expanded Newborn Screening Using Whole-Genome Sequencing. Pediatrics. 2016;137: S36–S46. doi:10.1542/peds.2015-3731H

58. Carlton J, Griffiths HJ, Horwood AM, Mazzone PP, Walker R, Simonsz HJ. Acceptability of childhood screening: a systematic narrative review. Public Health. 2021;193: 126–138. doi:10.1016/j.puhe.2021.02.005

59. Chambers D, Baxter S, Bastounis A, Jones K, Kundakci B, Cantrell A, et al. The acceptability of blood spot screening and genome sequencing in newborn screening: a systematic review examining evidence and frameworks. Health Technol Assess. 2025; 1–53. doi:10.3310/RTPQ2268

60. Liang NSY, Watts-Dickens A, Chitayat D, Babul-Hirji R, Chakraborty P, Hayeems RZ. Parental Preferences for Expanded Newborn Screening: What Are the Limits? Children. 2023;10: 1362. doi:10.3390/children10081362

61. Powell CM. Newborn Screening and Long-term Outcomes. Pediatrics. 2020;146. doi:10.1542/peds.2020-023663

62. Carlfjord S, Lindberg M, Bendtsen P, Nilsen P, Andersson A. Key factors influencing adoption of an innovation in primary health care: a qualitative study based on implementation theory. BMC Fam Pract. 2010;11: 60. doi:10.1186/1471-2296-11-60

63. Burton H, Adams M, Bunton R, Schröder-Bäck P. Developing Stakeholder Involvement for Introducing Public Health Genomics into Public Policy. Public Health Genomics. 2009;12: 11–19. doi:10.1159/000153426

64. Diepeveen S, Ling T, Suhrcke M, Roland M, Marteau TM. Public acceptability of government intervention to change health-related behaviours: a systematic review and narrative synthesis. BMC Public Health. 2013;13: 756. doi:10.1186/1471-2458-13-756

65. Lemke AA, Harris-Wai JN. Stakeholder engagement in policy development: challenges and opportunities for human genomics. Genet Med. 2015;17: 949–957. doi:10.1038/gim.2015.8

66. Schnabel-Besson E, Dikow N, Alex K, Mütze U, Straub H, Doll ES, et al. A multi-dimensional framework for establishing and managing a genomic newborn screening program. medRxiv; 2025. p. 2025.06.17.25329471. doi:10.1101/2025.06.17.25329471

67. Creswell JW, Clark VLP. Designing and Conducting Mixed Methods Research. ${number}nd édition. Los Angeles: SAGE Publications Inc; 2010.

68. Islam MR, Faruque CJ. Qualitative Research: Tools and Techniques. North Charleston, South Carolina: CreateSpace Independent Publishing Platform; 2016.

69. Sekhon M, Cartwright M, Francis JJ. Development of a theory-informed questionnaire to assess the acceptability of healthcare interventions. BMC Health Serv Res. 2022;22: 279. doi:10.1186/s12913-022-07577-3

70. Harpe SE. How to analyze Likert and other rating scale data. Curr Pharm Teach Learn. 2015;7: 836–850. doi:10.1016/j.cptl.2015.08.001

71. South L, Saffo D, Vitek O, Dunne C, Borkin M. Effective Use of Likert Scales in Visualization Evaluations: A Systematic Review. 2021 [cited 14 July 2025]. Available: https://osf.io/exbz8/

72. Norman G. Likert scales, levels of measurement and the “laws” of statistics. Adv Health Sci Educ. 2010;15: 625–632. doi:10.1007/s10459-010-9222-y

73. Koo M, Yang S-W. Likert-Type Scale. Encyclopedia. 2025;5: 18. doi:10.3390/encyclopedia5010018

74. Armstrong B, Christensen KD, Genetti CA, Parad RB, Robinson JO, Blout Zawatsky CL, et al. Parental Attitudes Toward Standard Newborn Screening and Newborn Genomic Sequencing: Findings From the BabySeq Study. Front Genet. 2022;13. Available: https://www.frontiersin.org/article/10.3389/fgene.2022.867371

75. White S, Mossfield T, Fleming J, Barlow-Stewart K, Ghedia S, Dickson R, et al. Expanding the Australian Newborn Blood Spot Screening Program using genomic sequencing: do we want it and are we ready? Eur J Hum Genet. 2023;31: 703–711. doi:10.1038/s41431-023-01311-1

76. Bombard Y, Miller FA, Hayeems RZ, Barg C, Cressman C, Carroll JC, et al. Public views on participating in newborn screening using genome sequencing. Eur J Hum Genet. 2014;22: 1248–1254. doi:10.1038/ejhg.2014.22

77. Fishbein M, Ajzen I. Belief, Attitude, Intention and Behavior: An Introduction to Theory and Research. Reading, Mass: Addison-Wesley; 1975.

78. Davis F. Perceived Usefulness, Perceived Ease of Use, and User Acceptance of Information Technology. Manag Inf Syst Q. 1989;13. Available: https://aisel.aisnet.org/misq/vol13/iss3/6

79. McCullough LB, Slashinski MJ, McGuire AL, Street RL, Eng CM, Gibbs RA, et al. Is Whole-Exome Sequencing an Ethically Disruptive Technology? Perspectives of Pediatric Oncologists and Parents of Pediatric Patients With Solid Tumors. Pediatr Blood Cancer. 2016;63: 511–515. doi:10.1002/pbc.25815

80. Pereira S, Robinson JO, Gutierrez AM, Petersen DK, Hsu RL, Lee CH, et al. Perceived Benefits, Risks, and Utility of Newborn Genomic Sequencing in the BabySeq Project. Pediatrics. 2019;143: S6–S13. doi:10.1542/peds.2018-1099C

81. Downie L, Halliday J, Lewis S, Lunke S, Lynch E, Martyn M, et al. Exome sequencing in newborns with congenital deafness as a model for genomic newborn screening: the Baby Beyond Hearing project. Genet Med. 2020;22: 937–944. doi:10.1038/s41436-019-0745-1

82. White AL, Boardman F, McNiven A, Locock L, Hinton L. Absorbing it all: A meta-ethnography of parents’ unfolding experiences of newborn screening. Soc Sci Med. 2021;287: 114367. doi:10.1016/j.socscimed.2021.114367

83. Hewlett J, Waisbren SE. A review of the psychosocial effects of false-positive results on parents and current communication practices in newborn screening. J Inherit Metab Dis. 2006;29: 677–682. doi:10.1007/s10545-006-0381-1

84. Tluczek A, Ersig AL, Lee S. Psychosocial Issues Related to Newborn Screening: A Systematic Review and Synthesis. Int J Neonatal Screen. 2022;8: 53. doi:10.3390/ijns8040053

85. Rueegg CS, Barben J, Hafen GM, Moeller A, Jurca M, Fingerhut R, et al. Newborn screening for cystic fibrosis — The parent perspective. J Cyst Fibros. 2016;15: 443–451. doi:10.1016/j.jcf.2015.12.003

86. Christie L, Wotton T, Bennetts B, Wiley V, Wilcken B, Rogers C, et al. Maternal attitudes to newborn screening for fragile X syndrome. Am J Med Genet A. 2013;161: 301–311. doi:10.1002/ajmg.a.35752

87. Blackwell K, Gelb MH, Grantham A, Spencer N, Webb C, West T. Family Attitudes regarding Newborn Screening for Krabbe Disease: Results from a Survey of Leukodystrophy Registries. Int J Neonatal Screen. 2020;6: 66. doi:10.3390/ijns6030066

88. Levenseller BL, Soucier DJ, Miller VA, Harris D, Conway L, Bernhardt BA. Stakeholders’ opinions on the implementation of pediatric whole exome sequencing: implications for informed consent. J Genet Couns. 2014;23: 552–565. doi:10.1007/s10897-013-9626-y

89. Persky S, Kaphingst KA, Condit CM, McBride CM. Assessing Hypothetical Scenario Methodology in Genetic Susceptibility Testing Analog Studies: A Quantitative Review. Genet Med Off J Am Coll Med Genet. 2007;9: 727–738. doi:10.1097/gim.0b013e318159a344

90. Genetti CA, Schwartz TS, Robinson JO, VanNoy GE, Petersen D, Pereira S, et al. Parental interest in genomic sequencing of newborns: enrollment experience from the BabySeq Project. Genet Med Off J Am Coll Med Genet. 2019;21: 622–630. doi:10.1038/s41436-018-0105-6

91. Dangouloff T, Hovhannesyan K, Mashhadizadeh D, Minner F, Mni M, Helou L, et al. Feasibility and Acceptability of a Newborn Screening Program Using Targeted Next-Generation Sequencing in One Maternity Hospital in Southern Belgium. Children. 2024;11: 926. doi:10.3390/children11080926

92. Ziegler A, Koval-Burt C, Kay DM, Suchy SF, Begtrup A, Langley KG, et al. Expanded Newborn Screening Using Genome Sequencing for Early Actionable Conditions. JAMA. 2025;333: 232. doi:10.1001/jama.2024.19662

93. Evans JR, Mathur A. The value of online surveys: a look back and a look ahead. Internet Res. 2018;28: 854–887. doi:10.1108/IntR-03-2018-0089

94. Ravalihasy A, Faye A, Diallo AI, Gaye I, Ridde V. A social acceptability scale: Validation in the context of government measures to curb the COVID-19 pandemic in Senegal. Ann Epidemiol. 2024;94: 49–63. doi:10.1016/j.annepidem.2024.04.004

95. Zeufack SCM, Omoto J, Owaya A, Adoyo E, Rop M, Osongo CO, et al. Feasibility of incorporating the Pocket colposcope into nurse-led cervical cancer screening programs in Western Kenya. Ecancermedicalscience. 2025;19: 1925. doi:10.3332/ecancer.2025.1925

96. Kiirya Y, Kitaka S, Kalyango J, Rujumba J, Obeng-Amoako GAO, Amollo M, et al. Acceptability of an online peer support group as a strategy to improve antiretroviral therapy adherence among young people in Kampala district, Uganda: qualitative findings. BMC Infect Dis. 2025;25: 461. doi:10.1186/s12879-025-10831-8

97. Mtenga AE, Maro RA, Dillip A, Msoka P, Emmanuel N, Ngowi K, et al. Acceptability of a Digital Adherence Tool Among Patients With Tuberculosis and Tuberculosis Care Providers in Kilimanjaro Region, Tanzania: Mixed Methods Study. Online J Public Health Inform. 2024;16: e51662. doi:10.2196/51662

98. Peay HL, Gwaltney AY, Moultrie R, Cope H, Boyea BL-, Porter KA, et al. Education and Consent for Population-Based DNA Screening: A Mixed-Methods Evaluation of the Early Check Newborn Screening Pilot Study. Front Genet. 2022;13: 891592. doi:10.3389/fgene.2022.891592

